# Assessment of glucocorticoid-induced enhancer activity of eSNP regions using STARR-seq reveals novel molecular mechanisms in psychiatric disorders

**DOI:** 10.1101/2022.05.18.22275090

**Authors:** Signe Penner-Goeke, Melissa Bothe, Nils Kappelmann, Peter Kreitmaier, Ezgi Kaya, Dorothee Pöhlchen, Anne Kühnel, Darina Czamara, BeCOME working group, Laura V. Glaser, Simone Roeh, Maik Ködel, Jose Monteserin-Garcia, Christine Rummel, Janine Arloth-Knauer, Laura Diener-Hölzl, Barbara Woelfel, Susann Sauer, Stephan Riesenberg, Michael J. Ziller, Marta Labeur, Sebastiaan H. Meijsing, Elisabeth B. Binder

## Abstract

Exposure to stressful events increases risk for psychiatric disorders. Mechanistic insight into genetic factors moderating the impact of stress can increase our understanding of disease processes. Here, we test 3662 SNPs from preselected expression quantitative trait loci in massively parallel reporter assays to identify genetic variants that modulate the activity of regulatory elements sensitive to glucocorticoids, important mediators of the stress response. Of the tested SNP sequences, 547 were located in glucocorticoid-responsive regulatory elements of which 233 showed allele-dependent activity. Transcripts regulated by these variants were enriched for those differentially expressed in psychiatric disorders in postmortem brain. Phenome-wide Mendelian randomization analysis in 4,439 phenotypes revealed potentially causal associations specifically in neuro-behavioral traits, including psychiatric disorders. Finally, functional gene scores derived from these variants were significantly associated with differences in physiological stress measures, suggesting that these may alter disease risk by moderating the individual set point of the stress response.

## Introduction

Trauma, chronic stress and exposure to adverse life events are among the most robust risk factors for a large range of psychiatric disorders.^1-3^ Accordingly, adverse life events or traumatic experiences substantially increase the risk for major depression, post-traumatic stress disorder, substance and alcohol use disorders, bipolar disorder, schizophrenia and other disorders.^4^ Despite their association with disease risk and disease presentation (more severe, more comorbidities, and more difficult to treat), we still have insufficient knowledge about the underlying molecular mechanisms, thereby limiting our ability to develop therapeutic approaches targeting the consequences of adversity exposure. Notably, not all exposed individuals develop disease, suggesting that underlying personal factors moderate the relationship between risk exposure and pathology.^5^ For example, additional exposure to protective or risk environmental factors, timing, and duration of the exposures have been suggested to moderate this relationship. Moreover, studies using SNP-based heritability measures in the UK Biobank cohort indicate a genetic component moderating adversity-associated psychiatric disorders with heritability estimates of 12% for major depressive disorder (MDD) alone, but of 24% in MDD patients with reported trauma.^6^ This supports findings of another study in an independent cohort demonstrating context-specific heritability in different complex traits, including gene x adversity effects in depression.^7^ Taken together, these studies suggest that genetic components moderate the risk for developing psychiatric disease following exposure to adversity.

Identifying specific genetic variants moderating the effects of adversity has proven difficult and the investigation of such interactions at the candidate gene, genome-wide and polygenic level have yielded inconsistent results.^8-13^ One approach is to uncover adversity-moderating genetic components directly from genome-wide interaction studies or other approaches using large cohorts. However, these approaches are hampered by statistical requirements for large sample sizes that come with increased heterogeneity due to differences and limitations in adversity definition and characterization, challenges of a comprehensive mapping of the exposome and differences in outcome definitions. An alternate approach to identify genetic variants that moderate the response to adversity is to dissect the genetic moderation of the more immediate impact of exposure to adversity, which elicits a complex multilevel physiological stress response. This response can be broken down into distinct biological components and offers the possibility to focus on specific potential biological mediator systems as well as on more proximal outcomes, such as stress-induced changes in gene expression.

One such component postulated to mediate the risk for psychiatric disorders is the stress hormone system^14 15^. Here, exposure to a threat or stressor leads to the activation of the hypothalamic-pituitary-adrenal (HPA)-axis, culminating in the systemic release of glucocorticoids (GCs) from the adrenal glands. GCs bind with high affinity to the mineralocorticoid receptor and with lower affinity to the glucocorticoid receptor (GR). Both are cytoplasmic receptors that translocate to the nucleus once activated and elicit a transcriptional response by binding to specific regulatory elements (REs) associated with target genes.^16^_6_ These transcriptional effects are essential for mediating a concerted stress response as well as for the negative feedback regulation of the HPA-axis once the stressor has subsided. Importantly, the dynamic regulation of the HPA-axis is altered in patients with different psychiatric disorders as well as in those with exposure to early/chronic adversity^17 18^ indicating that dysregulated GC activity is an important pathomechanism in adversity-related disease^15^. Genetic variants that alter GC-induced RE activity could thus be important moderators of the downstream, system-wide effects of stress hormones and hence disease risk. GC responsive REs, however, are not simply GR-binding sites given that less than 20% of the genomic loci occupied by GR exhibit GC-induced changes in RE activity in any given cell type.^19^

In this study, we used massively parallel reporter assays (MPRA) to identify genetic variants that specifically moderate GC-dependent RE function and then assess how such variants relate to disease risk and the physiological stress response **(see Fig. 1, graphical abstract**). To enhance the likelihood of identifying such variants, we screened all SNPs in 320 linkage disequilibrium blocks that we had previously shown to moderate GC-induced transcriptional responses in peripheral blood cells. ^20^

**Figure 1.**
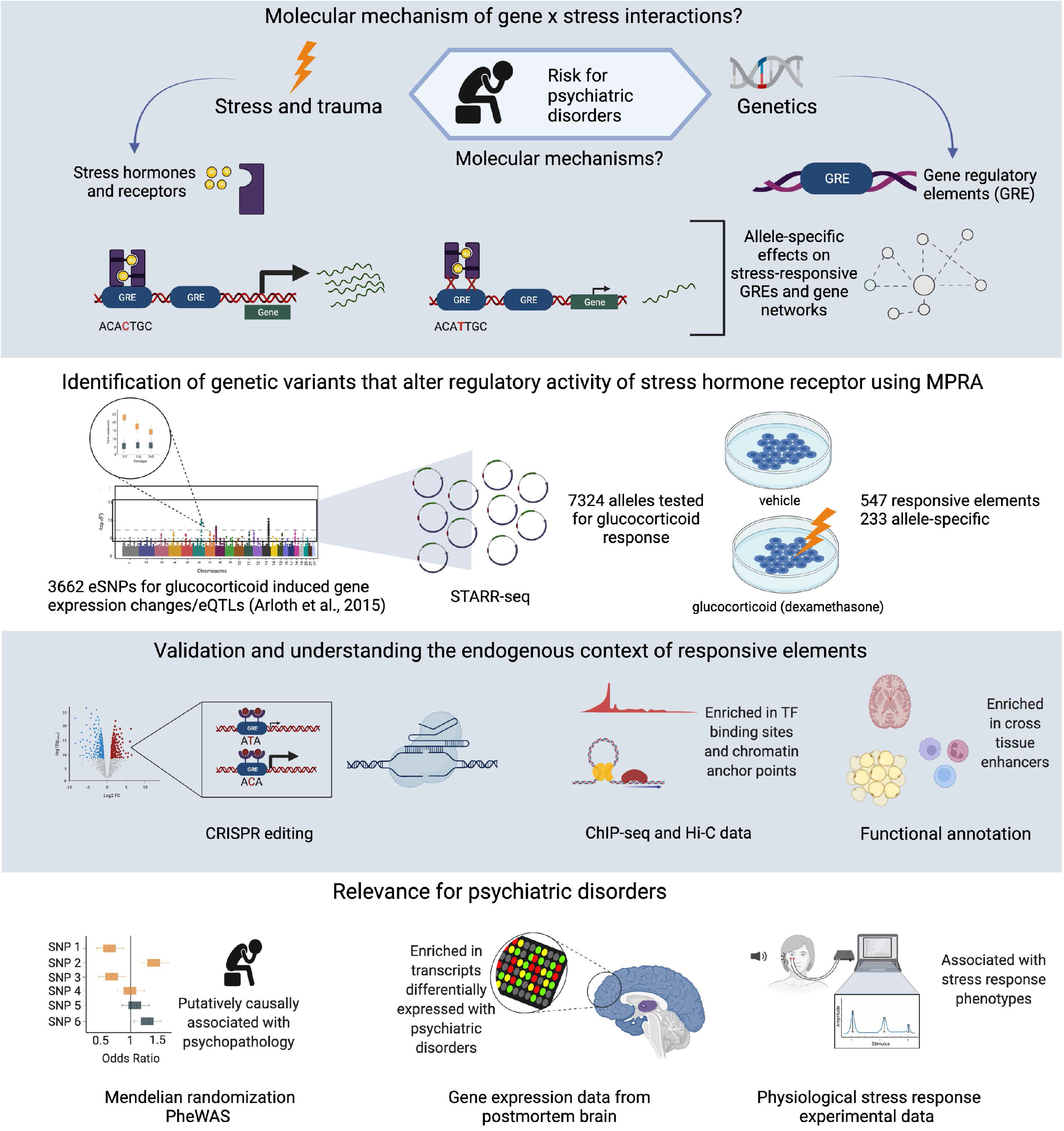
Graphical Abstract summarizing the rationale, design and outcomes of this study.

## Results

### Identification of functional regulatory elements modulating the response to stress in an allele-specific manner using STARR-seq

For our enhancer screen, we selected a set of 3,662 eSNPs **(Table S1)** that we had previously implicated in modulating the transcriptional response to the GR agonist dexamethasone (dex) by a genome-wide expression quantitative trait locus (eQTL) analysis (see **Fig. 2A**) and shown to be enriched among genetic variants associated with psychiatric disorders.^20^ These 3,662 GR-eSNPs were associated with 296 dex-responsive transcripts and organized into 320 independent genomic loci (eSNP bins), containing sets of eSNPs in linkage disequilibrium (LD; r^2^ >= 0.2). As previously reported,^20^ these eSNPs were enriched in enhancer regions, as defined by the Roadmap Epigenome Project, suggesting that modulation of target gene expression by these eSNPs could be mediated by variant-specific enhancer activities. In order to pin-point candidate causal variants among the linked eSNPs, we employed an MPRA variant, STARR-seq, to identify GC-responsive regulatory elements within the different eSNP loci. For the STARR-seq assay, candidate sequences are placed downstream of a minimal promoter, such that active enhancers induce their own expression and high-throughput sequencing reveals both the sequence identity and quantitative information regarding the activity of each sequence variant.^21^ We generated a reporter library containing 7,324 unique 201 bp oligonucleotides encompassing the 3,662 eSNP sequences as well as negative and positive control sequences (**Fig. 2B and methods**).^22^

**Figure 2.**
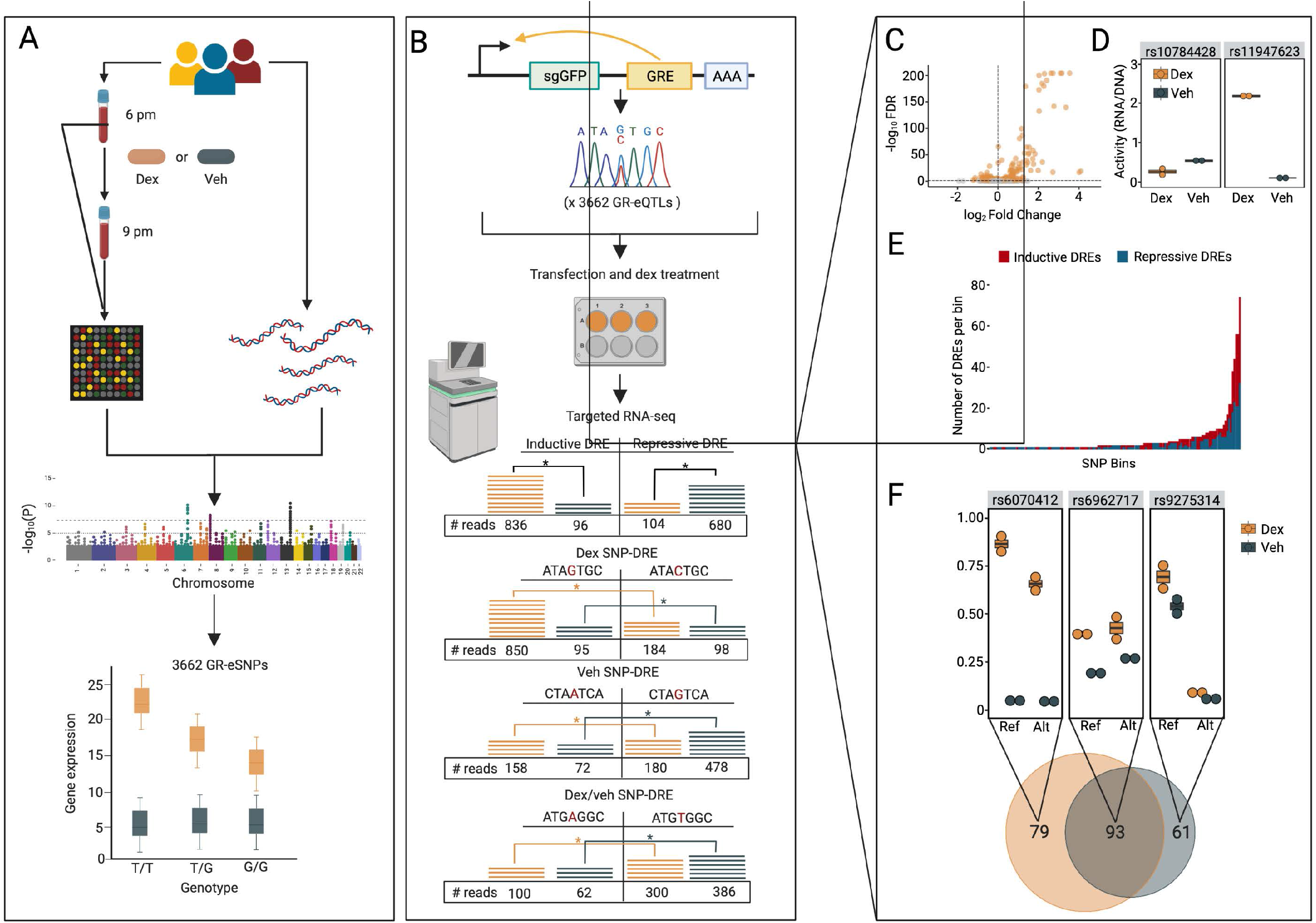
Identification of dexamethasone responsive regulatory elements using STARR-seq. **A**, Overview of the GR-eSNP analysis pipeline to identify dexamethasone (dex)-responsive eQTLs. Individuals were genotyped and their gene expression assayed before and 3 hours after oral administration of 1.5 mg dex resulting in the identification of 3,662 dex-responsive eQTLs (GR-eSNPs) as described in ^20^. **B**, Experimental procedure to assay the regulatory activity of each allelic variant of the 3,662 GR-eSNP regions using STARR-seq. For each GR-eSNP, we assayed a 201bp genomic region centered on the GR-eSNP: one with the reference allele, and the other with the alternative allele. U2OS-GR and U138MG cells were transfected with the STARR-seq library to quantify the activity of each variant for cells treated with dexamethasone or for cells treated with vehicle to assay their basal activity. **C**, Volcano plot showing the log^2^FC (dex/veh) and -log^10^ FDR for each of the variants assayed by STARR-seq. The 547 significant (FDR <0.1) dex-responsive regulatory elements are highlighted as orange dots. Inductive dex-responsive regulatory elements had a 1.8 greater mean fold change in activity in response to dex (mean log^2^FC = 0.59, SD = 0.64) compared to repressive dex-responsive regulatory elements (mean log^2^FC = -0.32, SD = 0.19). **D**, Regulatory activity for a representative inductive and repressive DRE is shown for both veh-(blue) and dex-treated (orange) conditions. **E**, Distribution of inductive and repressive DREs within the eSNP bins (median = 2 DREs/bin). **F**, Analysis of the STARR-seq data resulted in the identification of 79 DREs that exhibit allele-dependent activity exclusively in the dex condition, 61 exclusively in the veh condition, and 93 DREs with allele-dependent activity both in the veh and dex conditions. A representative example of each of these three types of variant-DREs is displayed. Activity of the enhancers are displayed as RNA/DNA ratios. The effect sizes of the SNPs on DRE activity were not significantly different between the veh and dex SNP-DREs, with an absolute mean fold change of 1.70 (SD = 0.56) and 1.59 (SD = 0.56), respectively.

These libraries were transfected into two cell lines that have previously been used successfully to study GC-related enhancer functions,^23,24^ an osteosarcoma cell line stably transfected with GR (U2OS-GR), and a brain glioblastoma cell line (U138MG) (**Fig. S1 A-B** for additional details on their GC response).

We then performed STARR-seq in these two cell lines in a vehicle (veh) condition and following stimulation with 100nM dex for 4 hours to identify active enhancers and changes in enhancer activity upon GC treatment including allele-specific differences (**Fig. 2B**). Across both cell lines and conditions, biological replicates were highly correlated (Pearson Correlation Coefficient >0.85, **Fig. S2**) and positive and negative controls behaved as expected, see methods. From this analysis (FDR <0.1) we identified 991 and 870 of the putative RE fragments as being active in the veh condition in the U2OS-GR and U138MG cells, respectively, for a total of 1071 unique REs. In the dex condition, 1023 and 872 of the putative REs were active in the U2OS-GR and U138MG cells, respectively. A combined analysis revealed that 508 and 66 sequences (for U2OS-GR and U138MG cells, respectively) changed their activity upon dex treatment and were termed dex-responsive regulatory elements (DREs) for a total of 547 unique DREs (**Fig. 2C-D, Table S2**). Among these, 51% showed increased activity after dex (inductive DREs) whereas 49% showed reduced activity after dex exposure (repressive DREs).

When mapping the DREs to the previously identified 320 eSNPs bins^20^, we found that 44% (N = 142) of the bins contained at least one DRE. Of these, 43% contained a single DRE, whereas the remaining 57% contained multiple DREs (median = 2, maximum = 69). Of the 81 multi-DRE bins, 12 exclusively contained inductive DREs, and 15 exclusively contained repressive DREs. The remaining 54 bins contained both repressive and inductive DREs residing within a single bin (**Fig. 2E)**, supporting previous observations that enhancers and silencers can interact within a genomic locus to facilitate fine-tuning of transcriptional regulation.^25^

In the identified 547 DREs, we next compared the STARR-seq activity of the reference to the alternative allele in the baseline (veh SNP-DREs) and dex condition (dex SNP-DREs) (**Fig. 2B**). We identified allele-dependent activity for 154 DREs (152 in the U2OS-GR and 2 in the U138MG) in the vehicle condition (veh SNP-DREs) and for 172 (164 in the U2OS-GR and 10 in the U138MG) in the dex condition (dex SNP-DREs), with 93 overlapping between the veh and dex conditions (dex/veh SNP-DREs) (**Fig. 2F, Table S3**). Veh SNP DREs show allele-specific differences in activity at baseline, while dex SNP DRE show allele-specific differences following stimulation with GCs (**Fig. 2B**).

### Functional annotation of the DREs

We identified 547 DREs of which 233 showed allele-dependent activity using STARR-seq. STARR-seq is an episomal and therefore ectopic reporter assay which assesses enhancer activity of DNA sequences regardless of their chromatin context by removing them from their endogenous chromatin context. ^21^ We thus sought to better understand the endogenous genomic context of these DREs. We began by examining whether GR binding is enriched at the DREs by intersecting these regions with data from GR ChIP-seq experiments performed in U2OS-GR cells ^26^. We observed that inductive DREs, but not repressive DREs, were enriched in GR binding sites (fold enrichment 1.9, permutation p-value = 0.03, for inductive DREs; fold enrichment = 0.81, permutation p-value = 0.84 for repressive DREs) (**Fig. 3A, 3B, 3C)**. However, overall most DREs lacked GR occupancy (491/508) likely reflecting their closed chromatin state in U2OS-GR cells, as the majority of GR binding occurs at open chromatin.^27^ Therefore, we also performed a motif analysis on the DRE sequences to identify enriched sequence motifs. Consistent with the ChIP colocalizations results, inductive DREs were enriched for the GR motif (FDR = 3.4 × 10^−11^) and the androgen receptor motif (FDR = 7.2 × 10^−8^), which has a highly similar motif to that of GR (**Table S4**). In contrast, and consistent with a lack of GR binding, the repressive DREs were not enriched in the GR binding motif (FDR = 0.13), but instead showed an enrichment of other TF motifs, including AP-1 (FDR = 5.6 × 10^−3^) and TEAD1 (FDR = 3.3 × 10^−3^), known tethering partners of GR.^28,29^

**Figure 3.**
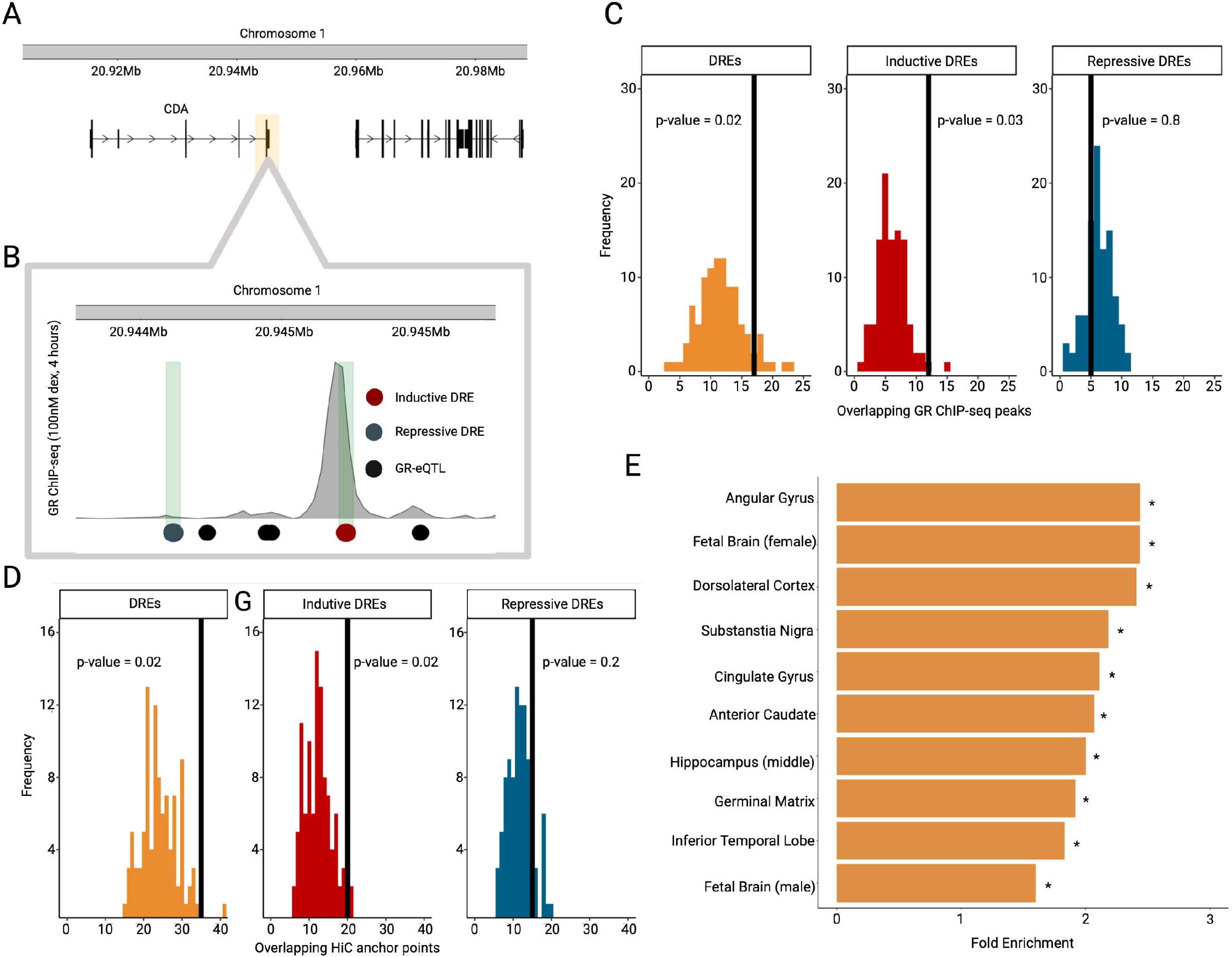
Understanding the endogenous context of the dexamethasone responsive regulatory elements. **A**, Representative genomic context of an eSNP bin (highlighted yellow box). **B**, Zoom-in of the eSNP bin showing GR ChIP-seq signal for the bin which contains GR-eSNPs (black dots), a repressive DRE (blue dot) and an inductive DRE (red dot) which overlaps with a GR ChIP-seq peak. **C**, Distribution of overlapping GR-eSNPs and GR ChIP-seq peaks with vertical lines denoting the number of observed overlaps in the DREs (orange, p-value = 0.02), inductive DREs (red, p-value = 0.03), and repressive DREs (blue, p-value = 0.8). **D**, Distribution of overlapping GR-eSNPs and chromatin anchor points with vertical lines denoting the number of observed overlaps in the DREs (orange, p-value = 0.02), inductive DREs (red, p-value = 0.02), and repressive DREs (blue, p-value = 0.2). **E**, ChromHMM enrichment of DREs predicted to be located within enhancer regions across specific brain regions compared to randomly selected matched SNPs. * FDR < 0.05; ** FDR< 0.01; permutation FDR.

In addition to chromatin accessibility, the 3D chromatin architecture has also been shown to influence RE activity, with evidence that dex-induced GR binding increases chromatin interactions between REs and the transcription start site of the regulated transcripts via 3D chromatin loops.^30^ We thus evaluated the connection between DREs and long-range chromatin interactions by testing whether the DREs were enriched in chromatin anchor points using publicly available Hi-C data from U2OS-GR cells (**Fig. 3D**).^31^ The analysis revealed that overall, the U2OS-GR DREs (±5000 kb from center) were enriched in chromatin looping anchor points compared to all 3662 eSNPs (permutation p-value = 2 × 10^−2^, fold enrichment = 1.45). Again, this held true only for inductive (permutation p-value= 2 × 10^−2^, fold enrichment = 1.8) but not repressive (permutation p-value = 0.16, fold enrichment = 1.3) DREs.

Given that STARR-seq can assess enhancer activities of DNA sequences regardless of their endogenous chromatin context, we next examined whether the identified DREs were located within predicted enhancer regions in different tissues, including those relevant to neuropsychiatric disorders, such as brain. For this we annotated the DREs to ChromHMM states (18-state model) across multiple tissues and calculated fold enrichment using a background constructed from 100 randomly selected SNP bins of the same size.^32^ The analysis revealed that DREs were significantly enriched in predicted enhancers in tested (FDR <0.1) **(Fig. S3)**, including brain (fold-enrichment = 2.0, permutation-based FDR = 9.0 × 10^−3^) and DREs were enriched in enhancer regions of all ten (FDR <0.1) available brain regions **(Fig. 3E)**. This indicates that the identified DREs using STARR-seq in two cell lines also have cross-tissue regulatory function in native tissues.

Overall, these results indicate that the ectopic STARR-seq assay identifies active DREs that are enriched for hallmarks of active enhancers in tissues relevant for neuropsychiatric disorders, including specific TF binding sites, chromatin conformation, and histone marks.

### Validation of STARR-seq DREs

To validate the findings from the STARR-seq experiments, four DREs and three SNP-DREs were tested by individually cloning them into the STARR-seq vector and assessing expression levels using qPCR. Consistent with the findings of our screen, each of the DREs showed significant (p < 0.05) dex-responsiveness in the direction observed in the STARR-seq data **(Fig. S4B)**. Moreover, we could validate the allele-dependent activity for two out of three SNP-DREs **(Fig S4A)**. Furthermore, the magnitude of the log_2_ FCs of all individual REs from the validation assays were highly correlated to the magnitudes observed in the STARR-seq data (Pearson Correlation Coefficient of 0.98, p-value < 0.0001) (**Fig. 4A)** indicating that the STARR-seq results are reproducible and provide reliable quantitative information regarding enhancer activity, including for allele-specific analyses.

**Figure 4.**
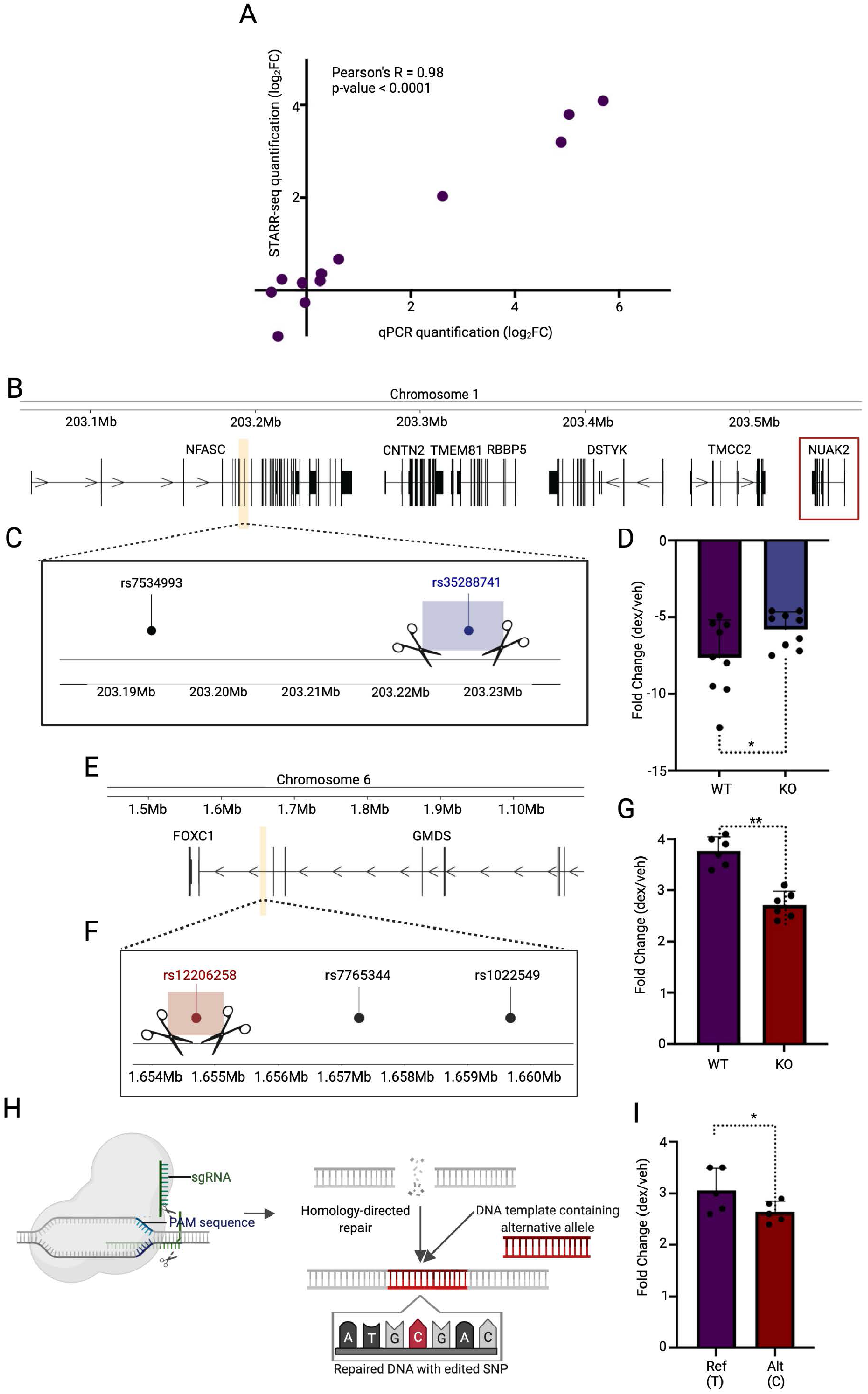
Exogenous and endogenous validation of the dexamethasone responsive regulatory elements. **A**, Correlation between the magnitude of log^2^ fold changes observed in the STARR-seq data with those observed in the qPCR validation of select candidate regions. **B-G**, To test the contribution of two dex SNP-DREs to the regulation of their predicted etranscripts, we removed each of the candidate enhancers in U2OS-GR cells and found that their deletion resulted in a blunted response to dex in a nearby gene. **B**, Schematic depicting the genomic locus containing an eSNP bin (highlighted orange) in relation to its predicted target gene, *NUAK2*, (red box) and **C**, a zoomed-in view of the region containing the repressive SNP-DRE rs35288741 with the CRISPR/Cas9-deleted region highlighted in blue. Black dot represents a flanking GR-eSNP not targeted. **D**, Deletion of the rs35288741 region resulted in a reduced repression of the *NUAK2* gene (p-value = 0.038, one-sided unpaired student t-test reported for all tests in this subheading). In addition to *NUAK2*, which is located ∼340kb from the deleted region, several other genes mapped to this region. Therefore, we also tested the effect of the rs35288741 deletion on five additional nearby genes. However, we found that among all the genes in the locus, deletion of the rs35288741 region had strong effects only on dex-regulated *NUAK2* expression except *DSTYK*, which was also significantly blunted **(Fig. S4C). E**, Schematic depicting the genomic locus containing an eSNP bin (highlighted orange) and its predicted target gene, *FOXC1* and **F**, a zoomed-in view of the targeted region containing the inductive dex SNP-DRE rs12206258, with the CRISPR/Cas9-deleted region highlighted in red. **G**, For *FOXC1*, activation was weaker when the rs35288741 region was deleted (p-value = 0.0024). To ensure the transcriptional changes observed in the KO cells were specific to the predicted target genes, we tested three canonical GR target genes (*FKBP5, SGK1, TSC22D3*) and did not observe differences in their dex-inducibility in the KO and WT cells, except for *TSC22D3* in the rs12206258 KO **(Figure S4 A-B)**, possibly due to *FOXC1’s* role as a transcription factor, which has been shown to bind at *TSC22D3*.^87^ **H**, Using homology-directed repair and a template, we edited SNP rs12206258. **I**, By generating homozygously edited (C/C) cells we observed a significant blunting of the transcriptional response to dex when compared to the homozygous wild type (T/T) cells (p-value = 0.040). As part of the genome editing strategy, we introduced an additional mutation to destroy the seed and prevent further cutting at the locus once the desired edit was made. To test if this additional change had an effect on *FOXC1* regulation, we also generated seed-mutated T/T cells and observed no difference between dex induction in these cells compared to the wildtype T/T cells demonstrating that the SNP-DRE, rs12206258, and not the seed mutation, is responsible for the blunted transcriptional response to dex. * p-value <0.05, ** p-value < 0.01.

To examine whether SNP-DREs identified in the episomal STARR-seq assay map to enhancers that are functional within the endogenous genomic context, we chose two dex SNP-DREs (one in an inductive DRE, the other in a repressive DRE) for CRISPR/Cas9 editing. Both SNPs were selected such that no other nearby SNPs were present, allowing a single SNP to be edited. The first SNP, rs35288741, is a repressive SNP-DREs located in the eSNP bin associated with transcriptional regulation of *NUAK2* (etranscript) **(Fig. 4B-C)**. The second SNP-DRE, rs12206258, is predicted to regulate the etranscript *FOXC1*, a gene induced in response to dex treatment (**Fig. 4E-F**). Both KOs showed significantly reduced dex-responsivity of their target genes, as expected (**Fig. 4D, G)** but generally not other dex-responsive transcripts, nor transcripts within the NUAK2 locus **(Fig S5A-D)**. Lastly, in order to test if it was indeed the specific allele change at rs12206258 in the deleted region that was driving the differential expression of the target gene, we edited rs12206258 with CRISPR/Cas9-SNP editing and evaluated the effect on *FOXC1* expression. Using homology-directed repair and a template harboring the C allele **(Fig. 4H)**, we generated homozygously edited (C/C) cells and observed a significant blunting of the transcriptional response to dex when compared to the homozygous wild type (T/T) cells (one-sided unpaired t-test, p-value = 0.040, **Fig. 4I)**.

### DRE-regulated transcripts are involved in brain function and psychopathology

We next sought to gain further insight regarding the connection between DRE and SNP-DREs and neuropsychiatric disorders by performing GO term enrichment analysis of associated etranscripts. The 3,662 GR-eSNPs that were identified in ^20^ are associated with the expression of 296 unique transcripts. Of these 296 transcripts, 122 (nearly 40%) are associated with DREs that we identified in our STARR-seq experiments **(Table S5)**. Of these 45 were associated with dex SNP-DREs, 36 with veh SNP-DREs, and 40 with dex/veh SNP-DREs. Functional annotation using GO term enrichment analysis of the 122 transcripts, revealed terms related to neural development as highly enriched (regulation of slits and ROBOs: adj.p-value = 1.66 × 10^−4^; axon guidance: adj.p value = 1.84 × 10^−4^; signalling by robo receptors: adj.p-value = 5.57 × 10^−4^). This enrichment of brain-related terms fits well to previous results, where GR-eSNPs transcripts were shown to be expressed in mouse brain and regulated by GR activation as well as social defeat stress.^20^ Further expanding the connection to expression in the brain, we find that DRE and dex SNP-DRE associated etranscripts are enriched in genes differentially expressed in post-mortem cerebral cortex of affected subjects across five neuropsychiatric disorders (SCZ, autism spectrum disorder (ASD), MDD, bipolar disorder (BPD), and alcohol abuse disorder (AAD))^33^ as compared to control subjects (DREs: FC = 4.2, permutation p-value = 0.01; dex SNP-DREs: FC = 4.1, permutation p-value =0.01). Notably, this was not the case with the veh SNP-DRE regulated transcripts (veh SNP-DREs FC = 1.05, permutation p-value = 0.44). These analyses support that the genetic variants that selectively alter the transcriptional response to stress hormones (i.e. dex SNP-DREs) could be of relevance to psychiatric disorders by altering brain gene expression following stress, especially as there was no enrichment etranscripts of SNPs in DREs that moderate only the baseline expression levels (i.e. veh SNP-DREs) among the differentially expressed genes.

### Dex SNP-DREs are enriched for psychiatric disease associated variants

Given that the eSNPs were previously found to be enriched for genetic variants associated with MDD and schizophrenia ^20^, an important question was whether the now identified subset of functionally validated SNPs moderating the GC-responsive enhancers (i.e. dex SNP-DREs) would show even stronger association with psychiatric disorders. For this, we compared the overlap of SNP-DREs (dex, veh and dex/veh SNP DREs) with variants nominally associated with psychiatric disorders (p-value ≤ 0.05) from a GWAS meta-analysis of eight psychiatric traits (BPD, MDD, attention deficit hyperactivity disorder, anorexia nervosa, obsessive compulsive disorder, SCZ, Tourette’s Syndrome, ASD)^34^ to the overlap of the eSNPs from the original selection. ^20^ Using 100 size-matched permuted eSNP sets as comparison, we found that dex SNP-DREs, but not the overlapping dex/veh, nor the veh SNP-DREs were enriched for variants associated with psychiatric disorders over the previously reported enrichment of the eSNPs ^20^ (dex SNP-DREs: fold enrichment = 1.5, permutation p-value = 0.03; dex/veh SNP DREs: 1.1, permutation p-value = 0.38; veh SNP-DREs: fold enrichment = 1.0, permutation p-value = 0.50) (**Fig. 5A)**. This indicates that disease association is more pronounced with the STARR-seq identified functional enhancer SNPs, than all eSNPs in the LD blocks.

**Figure 5.**
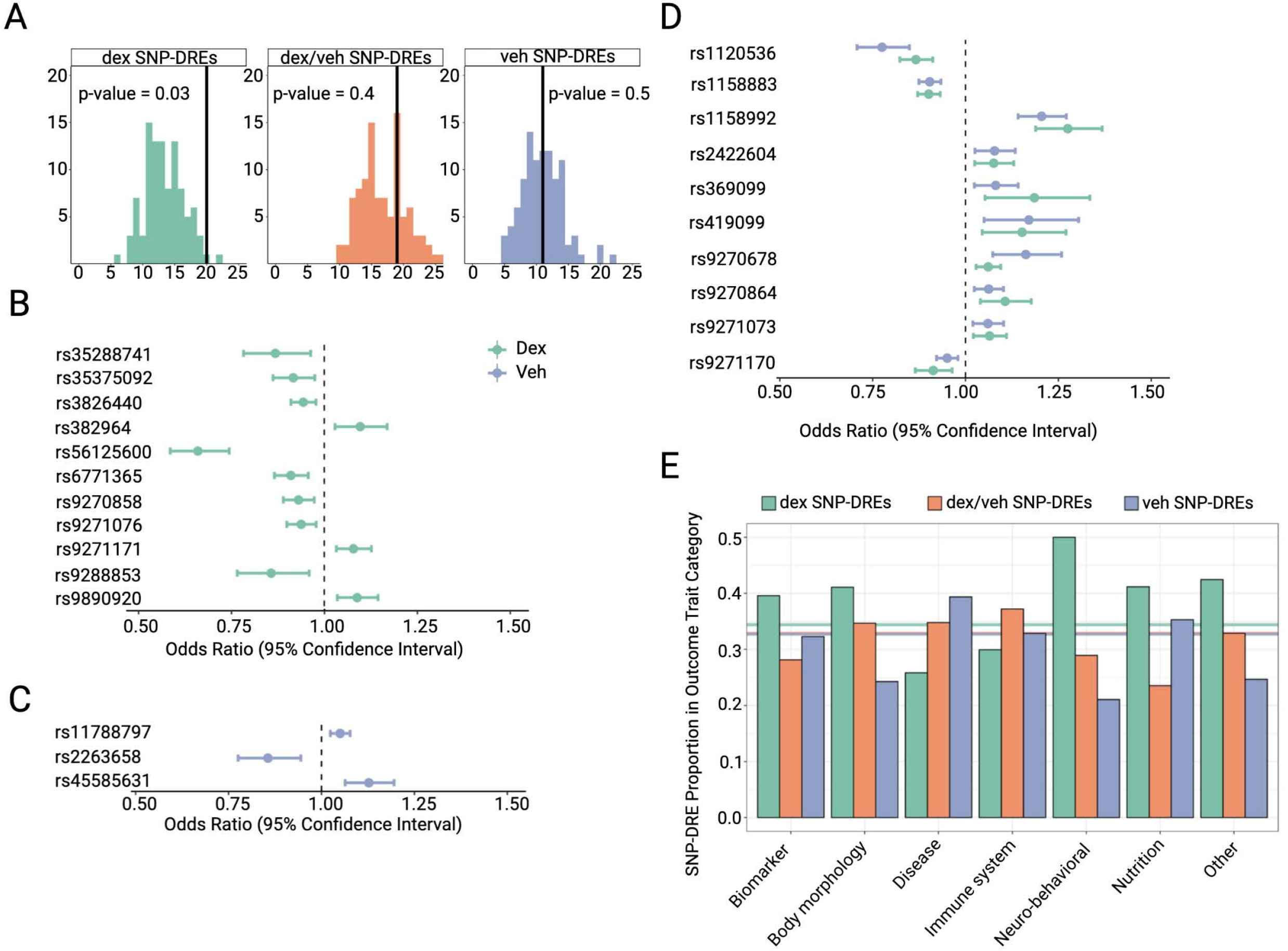
Dexamethasone responsive regulatory elements are relevant to psychiatric disorders. **A**, Enrichment analysis of dex (green), dex/veh (salmon), and veh (purple) SNP-DREs in GWAS hits (p value < 0.05) for cross disorder psychiatric risk compared to random GR-eSNPs sets of the same size. Vertical lines denote the number of observed overlaps between DRE/SNP-DRE and SNPs nominally significantly (p-value <0.05) associated with psychiatric disorders. Results from Mendelian Randomization to estimate the causal effect of **B**, dex SNP-DREs, **C**, veh SNP-DREs and **D**, dex/veh SNP-DREs on psychiatric disorders. **E**, Number of FDR corrected PheWAS associations per outcome trait category. Horizontal lines indicate the proportion of PheWAS association across all trait categories for dex (green), dex/veh (salmon), and veh (purple) SNP-DREs.

### Variant-REs are causally associated with psychiatric disorders

We next sought to determine whether the GC-effect moderating SNP-DREs we identified in this study could play a causal role in psychopathology. To test for causal relationships, we employed Mendelian Randomization (MR) using the exclusively veh (N = 61), exclusively dex (N = 79) as well as the dex/veh (N = 93) SNP-DREs as instruments. We weighed the allele-effect as the absolute log_2_FC of activity in the reference vs. alternative allele and associated the outcome with the psychiatric cross disorders GWAS.^34^ For veh SNP-DREs, we found only three (6.8% of 44 available for MR) with a significant (FDR < 0.05) putatively causal association with the psychiatric disorders. This number was markedly higher for dex SNP-DREs (eleven, 20% of 55 available), and also for dex/veh SNP-DREs (ten, 16% of 62 available) **(Fig. 5B-D, Table S6)**. As a control, we also performed MR on multiple non-psychiatric traits (height,^35^ Type 2 Diabetes,^36^ BMI,^37^ fracture risk,^38^ heart failure,^39^ chronic kidney disease,^40^ Crohn’s Disease^41^). With the exception of BMI and type 2 diabetes, we found that no other trait was causally associated with a dex SNP-DRE. In the cases of type 2 diabetes and BMI, three and nine (5.6% of 54 available and 26% of 34 available) dex SNP-DREs were causally associated, respectively. For the veh SNP-DREs, BMI, type 2 diabetes, fracture risk, and Crohn’s Disease all had significant MR results (BMI: 23% of 30 available; type 2 diabetes: 4.7% of 42 available; fracture risk: 3.3% of 30 available; and Crohn’s Disease: 1.9% of 53 available), indicating that the dex SNP-DREs are more specific to psychiatric traits than the veh SNP-DREs, which are enriched for various non-psychiatric disorders.

To examine whether the causal relationship to the dex SNP-DREs was overrepresented in psychiatric traits, we scaled up the MR analyses using a phenome-wide association study (PheWAS) approach. Specifically, we curated a list of 4,439 phenotypes from the MRC IEU OpenGWAS platform and applied MR separately on each outcome using a reduced list of 113 statistically independent (clumping threshold of R^2^= 0.001 using 10,000kb window) SNPs representing our 3 functional SNP groups (dex, dex/veh, and veh SNP-DREs). After correcting for the large number of (313,308) statistical tests, we observed 2,197 (0.7%) associations that passed our significance threshold (FDR <0.05). We categorised outcomes with significant associations into one of eight phenotype groups (biomarker, body morphology, disease, immune system, neuro-behavioral, nutrition, and other; **Table S7**). This revealed differing proportions of causally associated dex, dex/veh, and veh SNP-DREs across outcome trait categories (χ^2^(12) = 61.24, P<0.001). In fact, dex SNP-DREs had the highest proportion of significant MR associations in the neuro-behavioral trait category as compared to the seven other categories **(Fig. 5E, Table S8)**. Dex SNP-DREs accounted for 50% of all significant associations in this category, supporting a relative selectivity of these functional variants to neurobehavioral traits, the majority of which have also been reported to be affected by stress exposure. We list the SNP-DREs and FDR-corrected associations for the top exposures for each trait category in **Fig. S6**. To allow detailed exploration of our results, we created a dedicated Shiny app to assess PheWAS associations of individuals SNP-DREs (https://nprct.shinyapps.io/GRE_PheWAS/).

### Association between SNP-DREs and psychophysiological stress phenotypes

Given the enrichment of dex SNP-DRE in causal associations with neurobehavioral traits, we constructed a functional gene score (FGS) for these variants to test their association with variability in the physiological stress-response in a transdiagnostic, psychiatric, deep phenotyping study ^42^. The FGS was generated for 78 (of 79) of the dex SNP-DREs and weighted by the dex SNP-driven differential RE activity quantified by STARR-seq, as we did for the MR analyses **(Table S9)**. We focused on two widely-employed stress phenotypes: changes in salivary cortisol in response to a psychological stress task (available in N=183 ^43^), and the eyeblink startle reflex (available in N=171 ^44^), elicited by an abrupt and intense noise played during a fear-conditioning paradigm (**Fig. 6A, C**). We chose these phenotypes because both are affected in stress-related psychiatric disorders, such as anxiety,^44,45^ MDD,^46,47^ and PTSD.^48,49^ While not associated with cortisol levels before the stress task (p-value = 0.99), we found that the FGS was significantly positively correlated to changes in cortisol immediately after completing the task (beta = 0.34, p-value = 0.015), and that this association persisted 30 minutes post-task (beta = 0.26, p-value = 0.025) (**Fig. 6B**). In the eyeblink startle reflex, we observed that a higher FGS was associated with an increased startle magnitude (Beta = 0.20, p = 0.013), as well as a dampened startle habituation (Beta = 0.18, p = 0.031) (**Fig. 6D**). Taken together, our study shows that individuals with higher scores, i.e., more alleles with increased GC-regulatory strength, showed a higher and more prolonged cortisol response as well as a magnified startle reflex even after repeated stimulus presentation. Interestingly, both a heightened and prolonged cortisol response as well as increased startle amplitude and decreased startle habituation have been reported in patients with depression, anxiety disorders of post-traumatic stress disorder.^45-50^ These data provide preliminary evidence that the functional SNPs that modulate the transcriptomic response to stress also associate with difference in the stress response at the system-level.

**Figure 6.**
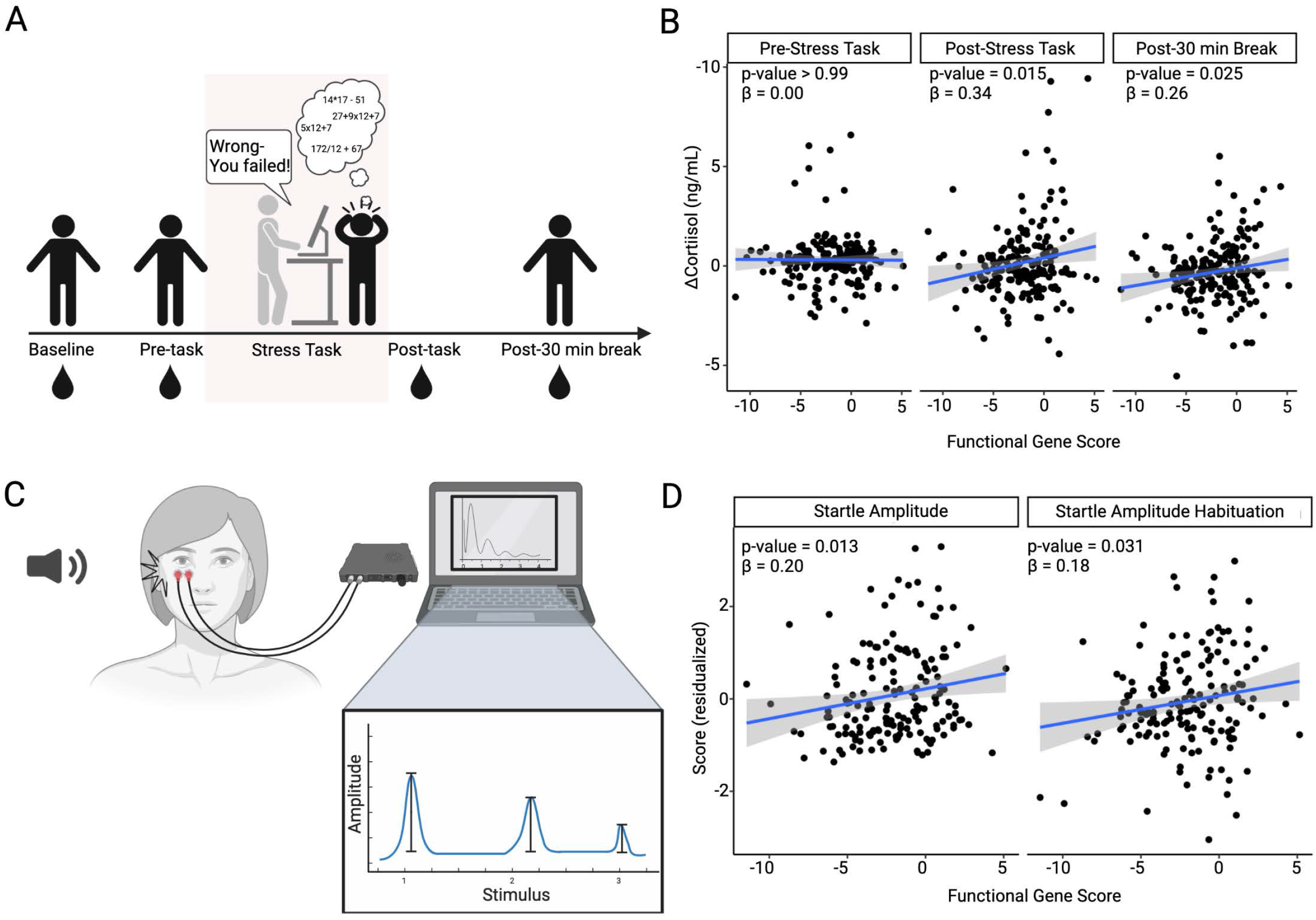
Dexamethasone responsive regulatory element modulate the physiological stress response. **A**, Experimental setup of a psychosocial stress task. Salivary cortisol was measured at baseline, prior to stress task, after completion of the task and following a 30-min rest period. **B**, Association of salivary cortisol changes compared to baseline with the functional gene score for the pre-test, post-test and post-break measurements. **C**, Experimental setup for a fear acquisition task eliciting an eyeblink startle response. **D**, Association between startle amplitude and habituation and the functional gene score. Note that an increase habituation z-score indicates a persisting startle response with multiple the startle probe.

## Discussion

Genetic variants relevant for common psychiatric disorders have been shown to be enriched in enhancer regions and to likely act in a cell-type and context dependent manner. MPRAs now allow us to map the cell-type and context specific effects of such regulatory variants.^51^ Using this method, we identified genetic variants that selectively modulate the activity of gene REs following stimulation by GCs to mimic a stress-context, an important risk factor for psychiatric disorders. Mendelian randomization analyses uncovered a causal relation between SNPs moderating GC-responsiveness (dex SNP-DREs) and risk for psychiatric disorders and neurobehavioral traits. While these variants were also causally associated with a number of other traits − including traits with well-documented links to GCs activity such as asthma, BMI and immune function ^52^ − neuro-behavioral traits showed the most pronounced over-representation for dex SNP-DREs. This suggests a relevant and specific contribution of genetic variants moderating the molecular response to stress hormones to risk for psychiatric disorders and traits. These SNP-DRE may thus provide a point of molecular convergence between environmental risk in the form of stress-induced increased GCs and genetic risk variants for psychiatric disorders.

While previous studies have identified GC-responsive REs using genome-wide STARR-seq,^53^_3_^53^_3_^53^ ours is the first to specifically explore such REs in the context of genetic variation. We focused on over 3,600 SNPs located within 320 individual LD bins that were identified as GR-eSNPs in a genome-wide analysis in peripheral blood. In the two GC-responsive cell lines we studied, we find 40% of these preselected bins to contain such DREs. Notably, the activity of DREs in the endogenous genomic context is often cell-type specific.^53^ This is partially due to differences in chromatin accessibility^27^ which plays a key role in dictating which genomic regions can be bound by GR. In addition, cell-type specific expression of transcription factors that act cooperatively with GR can influence the activity of DREs^27,54^ and therefore we expect that we would find more DREs by expanding our analysis to include additional cell types. Unfortunately, however, our efforts to establish STARR-seq in neural precursors or neurons in the 2D context failed due to a lack of robust GC-responsiveness (**Fig. S1C-E)**. Overall, balancing feasibility and using the right cell type is a major challenge in studies linking genetic variants with psychiatric disorders. Despite these limitations, our study indicates that testing MPRA in an episomal setting in cell lines picked for robust GC-responsiveness yields results of relevance for psychiatry given the enrichment of the identified DREs in predicted REs, including in brain (**Fig. 3E)**.

While 43% of the LD bins with a signal contain a single DRE, 57% have two or more, with up to 20 or more observed for specific bins **(Fig. 2E)**. In addition, we identify both inductive as well as repressive DREs, with a relatively equal distribution. While both inductive and repressive DREs are enriched for chromatin anchor points, only inductive DREs are enriched for GR-ChIP peaks. This is in line with previous studies showing that over 90% of the DREs mapping to GR-ChIP peaks were inductive.^19^ Consistent with the traditional model of GR repression that is independent of direct DNA binding, ^55^,^56^ we confirm that repressive DREs lack consensus steroid receptor-binding elements whereas they are enriched for the sequence motifs such as of AP1 (see Table S4) which is implicated in GR-dependent transcriptional repression.^28,29^ Of note, in close to 40%, we observe both repressive and inductive DREs in the same eQTL bin, supporting the notion of extensive regulatory fine tuning of the same target transcript.^25^ This fine-tuned genetic moderation of GC-induced transcriptional changes in brain tissue could be associated with disease risk, as etranscripts containing dex SNP-DREs in their eSNP bins were enriched among transcripts differentially expressed in cortex between individuals with psychiatric disorders and controls.^33^

Although MR has been more conventionally employed in the field of epidemiology to estimate the effect of a risk factor on an outcome variable using genetic variants that associate with the risk factor,^57^ in recent years it has also been applied to estimate causal effects of molecular QTLs on complex traits. For example, in psychiatric studies, MR analyses revealed a causal association between increased signaling of an inflammatory marker, IL-6, and suicidality,^58^ and glycemic traits with anorexia nervosa.^59^ Here, we decided to make use of SNPs with regulatory effects in DREs as instruments and quantitative molecular read-outs from the STARR-seq analysis as exposure variables to test for causal relationships with different traits using MR. Initial MR analyses revealed a preponderance of causal association of dex SNP-DRE with cross-disorder psychiatric risk as well as metabolic traits, but not others, such as height for example. The link to metabolic traits is not surprising, given the role of GR in glucose metabolism,^60^ and evidence for genetic coheritability between psychiatric traits (MDD, SCZ, AN) and BMI.^61,62^ PheWAS MR in over 4000 phenotypes confirmed that dex SNP-DRE were causally related to a large number of traits, but with a significant higher proportion of associations in neuro-behavioral traits, including psychiatric disorders than other trait categories.

These variants not only associate with disease risk, but a FGS computed for these dex SNP-DREs also correlates with features of the physiological stress response. Individuals with a higher FGS, i.e. more alleles with an enhanced response to GCs, showed an increased startle amplitude as well as decreased habituation to the startle reflex and also a heightened and prolonged cortisol response following a psychological stressor. These genetic variants may thus influence risk for psychiatric disorders by altering an individual’s physiological stress response set-point.

To our knowledge and despite the well-established link between exposure to stress and psychiatric disorders this is the first study to show that genetic variants modulating the transcriptomic response to stress may be causally involved in psychopathology, possibly by influencing the physiological stress response and stress-responsive brain transcription. Future studies need to expand the number of tested loci and diversify and refine the cell systems used for functional mapping.

## Online methods

### Cell culture

U138MG cells were purchased from the German Collection of Microorganisms and Cell Cultures GmbH. Cells were cultured in Minimum Essential Medium Eagle (Gibco) supplemented with 10% fetal bovine serum (Gibco), 1% sodium pyruvate (Gibco) and 1% antibiotic-antimycotic (Gibco). U2OS-GR cells ^63^ were cultured in Dulbecco’s Modified Eagle Medium with high glucose (Gibco) supplemented with 10% fetal bovine serum and 1% antibiotic-antimycotic. To activate GR, cells were treated with either 100nM dexamethasone (dex, Sigma Aldrich) or 0.001% ethanol as a vehicle control (veh) for 4 hours, unless stated otherwise.

### STARR-seq input library generation, sequencing, and STARR-seq

DNA fragments for integration into the human STARR-seq plasmid were designed by using the hg19 genomic coordinates of the GR-SNPs^20^ and extending them by 100bp up and down stream to generate 201bp fragments. All fragments had the following the following sequence composition:

‘5-**TAGAGCATGCACCGG**ACACTCTTTCCCTACACGACGCTCT—-*INSERT*—-AGATCGGAAGAGCACACGTCTGAACTCCAGTCAC**TCGACGAATTCGGCC**-3’.

Sequence homologous to the STARR reporter construct in bold; Sequence for p5 and p7 adaptors underlined.

Additionally, the following controls were included: known positive dex responsive REs defined by previous STARR-seq experiments in U2OS-GR cells (n=59)^64^ located near H3K27ac ChIP peaks, randomly selected genomic regions (n=52), and random sequences generated using a Markov Model (n=79). Of note, in the STARR-seq experiment (see below), we observed activation of 100% and 62.7% of the dex-inducible positive control regions included in our library in the dex condition whereas only 5.3 and 1.5% of our negative control regions were active in either the dex or veh conditions for the U2OS-GR and U138MG cells, respectively. An oligonucleotide pool containing all fragments and controls was ordered from Twist Biosciences and amplified using the Kappa HiFi HotStart ReadyMix PCR kit (Roche). The amplified fragments were cloned into the STARR-seq vector which was linearized with SalI-HF and AgeI-HF (New England Biolabs) at 37°C for 3 hours using In-Fusion HD Cloning Plus (Takara Bio) in ten parallel reactions and transformed into MegaX DH10B cells (Invitrogen). Libraries for sequencing were generated by PCR using 5 ng of purified plasmid DNA, 1.5 uL of NEB Universal Primer (New England Biolabs), 1.5uL of NEBNext Oligo 5 (New England Biolabs) and 25 uL of Kapa HiFi HotStart ReadyMix (Roche). The resultant plasmid library was sequenced on the Illumina MiSeq platform using the MiSeq Micro Reagent Kit V2 (Illumina, 300 cycles, 150 paired-end). The quality of the reads was assessed using FastQC and reads were trimmed using the Cutadapt tool^65^ to remove adaptor sequence. Reads were aligned to the ordered fragments using the bwa tool^66^ and only reads perfectly matching the reference were included in the analysis. The reads per fragment were summed and used for downstream normalization.

### STARR-seq

Five million cells U2OS-GR cells were transfected with 5ug of the input STARR-seq library using the Amaxa Nucleofector II Kit V and program X-001 (Lonza Bioscience). The next morning, the cells were treated with dex or the veh control for 4 hours. For U138MG cells, 5 million cells were plated in a 15 cm cell culture plate and transfected the next day with 15 ug of the STARR input library using Lipofectamine 3000 (Invitrogen) according to the manufacturer’s instructions. Eighteen hours following the transfection, cells were treated with dex or the veh control for 4 hours. After 4 hours of treatment, total RNA was collected using the RNeasy Midi Kit (Qiagen). mRNA was then isolated using the Dynabeads mRNA Purification Kit (ThermoFisher Scientific) and treated with TurboDNase (ThermoFisher Scientific). To generate cDNA, we used SuperScript III Reverse Transcriptase (ThermoFisher Scientific) using a primer to introduce random unique molecular identifiers to the cDNA (Table S10, primer 13). Furthermore, for each library, a unique index (Table S10, 15-26) and a universal adaptor (Table S10, primer 14) were used.

### Sequencing and data analysis

The STARR libraries were sequenced on the Illumina NovaSeq 6000 platform. Paired-end sequencing reads of 150bp were generated using the Novaseq 6000 SP Reagent Kit v1.5 (Illumina, 300 cycles). Sequencing quality was assessed using FastQC. The forward and reverse reads were stitched together using the tool, FLASH 1.2.11.^67^ Only stitched reads that had exactly 99 overlapping base pairs were included. After the forward and reverse reads were stitched together, UMI deduplication was performed using UMI-Tools^68^ for UMI-aware removal of PCR duplicates. The remaining reads were aligned and only reads that perfectly matched the sequences of the reference fragments were included. The fragments were filtered for a minimum read count of ten. For the downstream analysis, we used the R package MPRAnalyze 1.5.1^69^ that uses both the RNA count tables and the DNA plasmid library for normalization. Fragments were normalized for sequencing depth using the total sum scaling method (TSS). The normalized count tables were then used to determine which of the fragments were active REs. To this end, the negative control fragments included in our library were used to generate a null model. Then, using a generalized linear model, the transcriptional activity of each fragment was quantified using the function analyzeQuantification and the likelihood ratio test. Next, the REs with differential activity after dex treatment were identified using a comparative analysis (analyzeComparative). Lastly, regulatory elements with allele-dependent activity were identified using analyzeComparative Allele effects on DRE activity were quantified in each condition and those showing significant differential activity (FDR <0.1, Wald test) between alleles were classified as dex, veh, or dex/veh SNP-DREs, depending whether the allele-dependent activity was exclusive to the dex or veh condition, or was present in both.

### ChIP-seq

GR ChIP-seq sequencing data for the U2OS-GR cell line treated with 1uM dex for 90 minutes were publicly available from the sequencing read archive (Replicate 1: SRA accession SRX256867/SRX256891;82 Replicate 2: ArrayExpress accession E-MTAB-961683). Sequencing data were processed as described previously.^26^ To determine whether the DREs were enriched within in or near binding sites of the GR, a null distribution was generated by permutating the GR-eQTLs into 100 sets (with replacement) of the same size and counting the overlap with GR ChIP peaks.

### Motif Enrichment

Sequence motif analysis on the STARR-seq DREs was performed using the online tool, “TRAP (Transcription Factor Affinity Prediction)” available at http://trap.molgen.mpg.de/cgi-bin/home.cgi.^70^ The “multiple DNA sequence analysis” was performed using the Jaspar vertebrate database^71^ and human promoters as the background model.

### Hi-C analysis

Publicly available Hi-C data performed on thymidine synchronized U2OS (parental U2OS-GR cell line) cells. Data were acquired and processed as described previously.^72^ Only data generated from Hi-C experiments performed 360-minutes post-thymidine treatment were analyzed to ensure that the 3D structure had completely recovered from the treatment. A consensus interaction set was generated by creating a union set of regions from both biological replicates using the R package LoopRig 0.1.1. To determine whether the DREs were enriched within or near chromatin loops, a null distribution was generated by permutating the GR-eQTLs into 100 sets (with replacement) of the same size and counting the overlap with HiC anchor points.

### Annotation of DREs and variant-DREs with ChromHMM states

STARR-seq identified DREs were annotated using the R package HaploReg 4.0.2^73^ using the core 18-state model. Fifteen different broad tissue categories were predicted to assess cross-tissue functionality. To assess whether the DREs were predicted to be enhancers in tissues more relevant to psychiatric disorders, a sub-analysis was performed, which functionally annotated these variants in all available brain regions. To determine whether the DREs were enriched within enhancers, a null distribution was generated by permutating randomly selected SNPs (matched using SNPSnap ^74^ into 100 sets (with replacement) of the same size and quantifying how many were annotated as located within enhancer regions.

### Exogenous Validation

Individual fragments were ordered as “Gene Fragments” from Twist Biosciences. Identical to the fragments ordered for the STARR-seq experiment, the region of interest of each fragment was 201bp long. Each fragment was cloned individually into the linearized STARR-seq plasmid using In-Fusion HD Cloning (Takara Bio). Transformation was performed using Stellar Competent (Takara Bio) cells and the plasmids were isolated. The cloning products were sequenced using the Eurofins DNA Sanger sequencing service, using a single primer (Table S10, Primer 35). To test the activity of the cloned regions, two million U2OS-GR cells were transfected with 2ug of the STARR-seq vector containing the individual fragment using the Nucleofector IIB, Kit V and program X-001 (Lonza Bioscience). Sixteen to twenty hours following the transfection, the cells were stimulated with dex or the veh for 4 hours. RNA and DNA were extracted using the AllPrep RNA/DNA Extraction Kit according the instructions for adherent cell lines. cDNA was generated using the QuantiTect Reverse Transcriptase (Qiagen) kit following the manufacturer’s instructions except that two gene specific primers, GFP and RPL19 (Table S10 Primers 29-30) replaced the random hexamers. Regulatory elements activity was assessed with qPCR using primers for *RPL19* and *GFP* (Table S10, primers 31-34).

### Endogenous Validation

#### CRISPR genome editing for generation of knockout U2OS-GR cells

CRISPR-Cas9 genome editing was used to create genomic deletions of the regulatory elements harboring the variant rs12206258 and rs35288741 controlling *FOXC1* and *NUAK2* expression, respectively, according to the Horizon Discovery protocol (https://horizondiscovery.com/-/media/Files/Horizon/resources/Protocols/cas9-protein-nucleofection-protocol) with some modifications. gRNAs pairs (crRNA/tracrRNA duplex, IDT) and recombinant spCas9 protein (IDT) were selected to cut approximately 400 bp upstream and downstream from the desired SNP (Table S10, 36-39). crRNAs were designed using the Benchling webtool (https://benchling.com). To delete the region of interest, one million U2OS-GR cells were transfected by electroporation according to the manufacturer’s instructions in 100uL of nucleofection buffer (Nucleofector Kit V, Lonza Bioscience), 300 pmol electroporation enhancer (IDT), 300 pmol of each gRNA, and 100pmol Cas9 (IDT).

Subsequently, edited cells were plated in one well of a 6-well plate for analysis and further propagation. Two days post-electroporation, 80% of the bulk cells were expanded and 10% was used for bulk genotype analysis. The other 10% of the cells were plated in a 10 cm dish to generate single-cell-derived clonal cell lines. After seven days, colonies were picked for propagation and DNA isolation. DNA extraction was done using 30 µl QuickExtract DNA Extraction Solution (Lucigen). Briefly, bulk cells and clonal cell lines were dissociated, pelleted, resuspended in the extraction solution and incubated at 65 °C for 10 min and finally 98 °C for 5 min. PCR was done using the Q5 high fidelity master mix (New England Biolabs) and 4.5 µl of cell extract in a total volume of 10 µl. The thermal cycling profile of the PCR was: 98 °C 30 s; 30 × (98° 10 s, 64 °C 10 s, 72 °C 50 s); 72 °C 2 min. Primers (Table S10, Primers 40-43) were ordered from Sigma-Aldrich. Automated electrophoresis technique (DNA screen tape analysis, Agilent) and Sanger sequencing (Eurofins) were used to confirm the presence of CRISPR/Cas9-mediated knockout mutants in the bulk population. RT-qPCR for gene expression analysis of predicted target genes were performed in a vehicle and dex condition (Table S10, 44-49) and using YWHAZ for normalization. Given the high knockout efficiency for the rs35288741 region, the gene expression analysis was performed on the bulk population. For the rs12206258 the efficiency was lower and single-cell derived clonal lines homozygously edited for the region of interest were used.

#### SNP specific genome editing

CRISPR-Cas9 mediated single nucleotide editing was performed for the rs12206258 controlling FOXC1 expression. The 200 bp HDR templates (Table S10, Primers 50-51) were synthesized as duplex oligonucleotides by IDT. A point mutation outside the GR binding site that changes G into T (depicted in a bold letter), was introduced in the template to avoid repeated Cas9-editing modifications. gRNA (5’_GATCATGTTCCCTGAGCAGC_3’, PAM TGG) for genome editing was designed using the CHOPCHOP webtool (https://chopchop.cbu.uib.no/) and cloned into the sgRNA/Cas9 expression construct PX459 (GenScript). To generate clonal cell lines with the HDR-induced sequence changes, two million U2OS-GR cells were transfected using 4 μg of the gRNA construct and 200 pmol of the HDR template by nucleofection (Lonza Nucleofector kit V) according to the manufacturer’s instructions. Subsequently, successfully transfected cells were selected by treating cells with puromycin (5 μg/ml) for 48h. To increase gene editing by HDR, cells were treated with 2 μM M3814 NHEJ inhibitor (MedChem Express) for 3 days. Single-cell– derived clonal cell lines were genotyped by PCR using genomic DNA isolated with QuickExtract DNA Extraction Solution (Lucigen) and primers binding outside the HDR template (Table S10, Primers 52-53). Homozygous cells for rs12206258 (reference allele), the alternative allele and control cells (homozygous for rs12206258 without point mutation) were cultured and treated with vehicle or dexamethasone (100nM) for 4 hours. RNA was extracted (RNA Extraction Kit, Lexogen), reverse transcribed and analyzed by qPCR for gene expression analysis of *FOXC1* and *YWHAZ I* (Table S10, Primers 44-49).

#### Assessment of KO cell lines using qPCR

RNA was extracted from KO cell lines treated with dex and a veh control using the RNeasy Kit (Qiagen) and cDNA was generated using the Quantitect Reverse Transcriptase kit according to manufacturer’s instructions. (Qiagen). RT-qPCR for gene expression analysis of GR target genes and genes surrounding the *NUAK2* locus were performed in a vehicle and dex condition using PrimeTime qPCR assays (IDT, Table S10, 1-4, 54-60) and using *YWHAZ* for normalization, and Prime Time Gene Expression Master Mix according to the manufacturer’s instructions (IDT).

#### Enrichment of transcripts differentially expressed in psychiatric disorders

An enrichment analysis was performed on the DRE-associated transcripts as identified in a previous GR-stimulated eQTL study^20^ to determine whether they were overrepresented among transcripts shown to be differentially regulated in the prefrontal cortex of subjects with psychiatric disorders.^33^ All DREs, dex-DREs and veh SNP-DREs associated transcripts were tested to detect whether the subsets were differentially enriched. To this end, a randomly selected background set of transcripts from Illumina_humanht_12_v3 (the array used in Arloth et al.,) were permuted into sets (of the same size as the DRE-subset of interest) and overlapped transcripts differentially regulated in the prefrontal cortex of subjects with psychiatric disorders ^33^. The number of overlapping transcripts was counted. This was performed 100 times to generate a null distribution and compared to the number of overlapping transcripts differentially regulated in psychiatric disorders.

#### Mendelian randomization on single phenotypes

To test for a potentially causal effect of the differential DRE activity (driven by the variant-DREs) on psychiatric traits, a two-sample MR approach was employed using the R package “TwoSampleMR” ^75,76^. The instrumental variable was set as all variant-DREs and the log-fold change between the two alleles was used as the beta estimate. The exposure variables were the corresponding DREs showing allele-dependent activity, and the GWAS summary statistics for the PGC cross-disorder variants were used to define the outcome variables. An FDR threshold of <0.05 was used to define whether the SNP-DREs had a significant causal effect on psychiatric traits.

### Phenome-wide association study (PheWAS) analyses

#### Selection of genetic instruments

As exposure phenotypes, we used effect and variance estimates of identified SNP-DRE alleles that were functionally classified as dex, dex-veh or veh SNP-DREs. These SNPs were grouped according to their associated gene (or multiple genes if applicable) and, in order to retain statistically independent SNPs as genetic instruments, clumped separately for dex, dex-veh and veh SNP-DREs using a 10,000kB window and R^2^=0.001 according to a European reference population. This procedure led to retention of one single SNP-DRE per gene for the final analyses due to high LD between SNPs associated to the same gene.

To create the Shiny app, data preparation and analyses were repeated using the same approach, but without the clumping procedure. This choice of separating the analyses with and without clumping was made to allow not only multiple comparison correction of statistically independent Mendelian randomization tests and category-based comparisons for main analyses but also full browsability of individual SNP-DREs.

#### Outcome Phenotype Selection

As outcome phenotypes, we selected the PGC cross-disorder phenotype obtained from a GWAS of 232,964 cases with one of eight psychiatric disorders (anorexia nervosa, attention-deficit/hyper-activity disorder, autism spectrum disorder, bipolar disorder, major depression, obsessive-compulsive disorder, schizophrenia, and Tourette syndrome) compared to 494,162 healthy controls.^34^ In addition to this theoretically selected outcome phenotype, we selected a large number of phenotypes that were available on the MRC IEU OpenGWAS platform.^77^ The IEU OpenGWAS platform categorizes phenotypes into different phenotype batches and we initially selected 5,650 phenotypes originating from the UK Biobank study and the NHGRI-EBI GWAS Catalog (see Table S7 for trait category annotation of 698 phenotypes with significant associations).

From this original phenotype list, phenotypes were filtered using a three-step procedure. First, GWAS in non-European populations were removed. Second, duplicated phenotypes were removed in a semi-automated fashion: Duplicate phenotypes with identical names were removed automatically and phenotypes with high similarity in trait names (quantified using the restricted Damerau-Levenshtein distance with threshold >0.8 implemented in the stringdist package)^78^ were manually screened and removed. In each case, the duplicated phenotype originating from the GWAS with larger sample size was retained. Third, we manually excluded phenotypes not of interest to this study such as “Patient Care Technician responsible for patient data” or “Day-of-week questionnaire completion requested”. This procedure resulted in a final list of 4,439 phenotypes combined with the cross-disorder phenotype.

#### Mendelian randomisation (MR) analyses

MR analyses were conducted using the TwoSampleMR package.^79^

Variant data from exposure and outcome phenotypes were harmonised by trying to infer positive strand alleles and removing ambiguous and palindromic SNPs. MR analyses were then applied to all remaining exposure-outcome pairs using Wald ratio estimation as the method of choice for single SNP MR.^80^ This procedure resulted in a large number of 313,308 MR association estimates for all exposure-outcome combinations with available SNP data. We corrected for multiple testing using the Benjamini-Hochberg method.^81^ For the Shiny app and resulting MR analyses on all possible exposure-outcome associations without clumping, 829,053 MR association estimates were retrieved, which were also corrected using the Benjamini-Hochberg method. To interpret results from these analyses, all phenotypes with at least one FDR-significant association were grouped into the following seven categories: Biomarker, body morphology, disease, immune system, neuro-behavioral, nutrition, and other (Table S8). Using these categories, we applied a χ2 test to assess deviations from expected proportions of dex, dex-veh, and veh SNP-DREs per phenotype category. We also extracted the SNP-DREs with the most PheWAS associations in each of these categories as well as the outcomes that displayed associations with the largest number of SNP-DREs.

#### Associations between functional gene risk score and stress- and threat related endophenotypes

Next, we tested whether a functional gene score derived from the dex SNP-DREs predicts the response to stress or threat in a dimensional sample of healthy controls and patients with mood and anxiety disorders (BeCOME-study^42^ registered at ClinicalTrials.gov : NCT03984084). The Ethics committee of the Ludwigs Maximilians University gave ethical approval for this work and all participants gave an informed consent. All participants underwent in depth genotyping and phenotyping and completed two days of inhouse measurements, including a psychosocial stress task on the second day (N=187) as well as a classical fear-conditioning task on the first day (N=171). The cortisol response to stress and the startle response during fear acquisition were used as stress and threat-related endophenotypes. We then predicted those endophenotypes based on the functional gene risk scores using linear models including the covariates age, sex, and diagnosis of a mood or anxiety disorder within the last 12-months.

#### Calculation of functional gene risk score

Data from all participants was genotyped with the Illumina Global Screening array. After genotyping, we performed a stringent quality control in PLINK: we removed SNPs if they presented with a callrate below 0.98, with a minor allele frequency below 0.01 or if they were out of Hardy-Weinberg-Equilibrium (p<1.0e-05). Furthermore, we excluded individuals with a call rate below 0.98 as well as individuals who were MDS outliers (presenting with a position which was located more than four standard deviations from the mean on any of the first ten MDS axis of the IBD matrix) or outliers in heterozygosity (presenting with a heterozygosity more than 4 standard deviations from the mean heterozygosity). Furthermore, if siblings were present (as measured by pihat (i.e., the fraction of identical genotypes) > 0.125), only one random individual from each pair was kept in the analysis. After quality control, 386,848 SNPs were put forward to genotype imputation of missing SNPs. Phasing and imputation was performed using shapeit2 and impute2 using the 1,000 Genomes Phase III sample as reference. After imputation, we ran an additional round of quality control, removing SNPs with info score below 0.6, minor allele frequency below 0.01 or out of Hardy-Weinberg-Equilibrium (p<1.0e-05). The final imputed dataset comprised 9,651,000 SNPs. Of the 79 SNPs that showed a fold-change after dex but not after vehicle stimulation, 78 SNPs were mappable in the BeCOME sample (73 SNPs were directly available, 5 SNPs were included via proxy SNPs (LD r2 to original SNP > .80, based on 1000 Genomes European sample). The log fold change of these 78 SNPs was used as the effect size measure to calculate functional gene risk scores, based on the imputed dosages, using PLINK (see Table S10 for an overview of the included SNPs and their associated effect sizes).

#### Stress task and cortisol measurement

fMRI and cortisol samples were acquired during the psychosocial stress task in the BeCOME study.^43^ To measure baseline cortisol the first saliva sample was taken upon arrival (T1, n = 197). Approximately 20 minutes later before entering the scanner and after placement of an intravenous catheter for additional blood sampling in 73 participants (39%), a second sample (T2, n = 197) was taken. Stress was induced by performing arithmetic tasks under time pressure and with negative feedback^43^. After completion of the task and approximately 10 minutes after the stress condition, another saliva sample was taken (T6, N=187). This was followed by a 30-min rest period and a concluding resting-state scan before a last saliva sample (T8, N =183) was taken to measure recovery of the cortisol response. The cortisol stress response was calculated as change in salivary cortisol (ΔCort) between baseline (T1) and the end of the task (T6) or the end of the post-task resting period (T8), respectively. To account for potential pre-task cortisol responses induced by the blood sampling procedure in a subset of participants, we included an additional covariate (dummy-coded) classifying participants with a cortisol response (ΔCort > .91 ng/ml ^43^) 20 min after the placement of the IV (T1) as pre-task responders in the regression models.

#### Fear acquisition task and startle response quantification

Fear acquisition consisted of three blocks where participants (N = 171) learnt to associate three distinct geometric shapes with either no aversive outcome, an airblast, or an electrical shock.^44^ In each block, each stimulus type was presented four times in a pseudo-randomized order. Aversive outcomes occurred in 75% of trials. Startle probes were delivered during the presentation of the geometric shapes in 75% of trials and in 40% of inter-trial-intervals. Startle probes consisted of 40 ms white noise at 108 dB with near-instantaneous rise time. Startle amplitude was quantified as the difference between the baseline and the maximum muscular response in a window of interest from 20 to 120 ms after the startle probe. Startle peak latency was defined as time from sound onset to the identified peak. We averaged the responses for each of the stimulus types across the three fear acquisition blocks resulting in 12 amplitude and 12 latency variables for fear acquisition. We additionally calculated four startle habituation indices for amplitude and latency respectively. In order to lower the number of variables and to avoid multicollinearity, we reduced amplitude, latency and habituation measures of the startle response to stable components using principal component analysis (PCA) and the Varimax rotation in SPSS (Version 26, IBM). The PCA resulted in four components representing amplitude (PC1), latency (PC2), latency habituation (PC3) and amplitude habituation (PC4). For a more detailed description of the task and readout quantification please refer to^44^.

## Methods related to supplemental Figure 1

### Generation of iPSCs

Two induced pluripotent stem cell (iPSC) lines were generated as previously described ^82^ from 2 distinct healthy donors. Cells were quality controlled by immunofluorescence for OCT4, NANOG, and SOX. Additionally, they were analysed for their ability to form embryoid bodies expressing markers of all 3 germ layers. iPSC were cultivated on matrigel (Corning, 1:100 in DMEM/F12) coated plates in in StemMACS™ iPS-Brew XF (Miltenyi Biotech). Cells were passaged twice a week with 1:4 accutase (Merck) in PBS (ThermoFisher), centrifuged in DMEM/F12 (ThermoFisher Scientific) at 300 x g for 5 min. IPSCs were replated in iPS Brew supplemented with 1x RevitaCell (RC, A2644501, ThermoFisher Scientific). For counting purposes (LUNA-II™, Logos Biosystems), iPSC were dissociated with 1x accutase and spun down in DMEM/F12 containing 1% BSA (11020021, ThermoFisher Scientific).

### iPSC derived neural precursor cells (NPCs)

Neural progenitor cells (NPCs) were differentiated from iPSCs according to a slightly modified protocol ^83^. After the 1x accutase dissociation, cells (200,000 cells/cm^2^) were plated on matrigel (1:30) coated 6well plates in StemMACS iPS-Brew XF medium supplemented with RevitaCell™. On the next day (day 0) the media was changed to KSR medium containing 41 ml of Knockout DMEM, 7.5 ml Knockout Serum Replacement, 1 mM L-glutamine, 0.1 mM MEM-NEAA, and 0.1 mM β-mercaptoethanol (all ThermoFisher Scientific) including small molecules LDN193189 (250 nM, Miltenyi Biotech), SB431542 (10 µM, Miltenyi Biotech), XAV939 (5 µM, Tocris), PD0325901 (1 µM, Miltenyi Biotech), SU5402 (5 µM, Tocris), DAPT (10 µM, Miltenyi Biotech). LSB+XAV were added from day 0-8, and PD/SU/DAPT were added between day 2-8. 1/3 N2/B27 (ThermoFisher Scientific) for day 4 and 5, 2/3 N2/B27 for day 6, 7 and 8 were additionally added to the media. B27 was depleted for 24h before GR stimulation in day-8 media. Each sample consisted of one well of a 12 well plate.

### iPS derived induced neurons

iPSCs were infected in iPS Brew with 1x RevitaCell and 6 µg/ml polybrene (Sigma Aldrich) with 3 - 15 µl virus (FUW-M2rtTA, Addgene and pTet-O-Ngn2-puro, Addgene) coding for the TET-ON system (reverse tetracycline-controlled transactivator and TET responsive promoter), mouse Ngn2, and puromycin resistance gene, respectively ^84^. 500,000 cells / ml were incubated with the virus at RT for 10 min and plated at a density of 2.5 mil on a 10 cm dish coated with matrigel. Ngn2 expression was induced by doxycycline (2 µg/ml) to the differentiation media from day-1 until day-21. Cells were forebrain patterned with 2 µM XAV939, 10 µM SB431542, 100 nM LDN193189 in KO-DMEM supplemented with 15% KO-Serum, 1x GlutaMax, 50 µM 2-mercaptoethanol on day-1. On day-2, cells were incubated with a 1:1 mixture of the media from day-1 and day-3. On day 3, cells were treated with DMEM/F12, 1x N2, 1x GlutaMax, 1x MEM-NEAA, and 10 µg/ml puromycin. On day-4, 80,000 cells / 48-well were seeded in Neurobasal medium containing doxycycline (2 µg/ml), 1x GlutaMAX (ThermoFisher Scientific), 1x B-27 (no Vitamin A, ThermoFisher Scientific), 1x MEM-NEAA (ThermoFisher Scientific), BDNF (Miltenyi Biotech), GDNF (Miltenyi Biotech), CNTF (Miltenyi Biotech) on previously coated neuronal differentiation plates: 12.5 µg/ml Poly-L-ornithine in DPBS O/N at 37 °C, 3 washes with DPBS, laminin (1 µg/ml) and fibronectin (2 µg/ml) in DPBS. Primary mouse astrocytes were added at day-7 at 5,000 / 0.95 cm^2^ to further mature the differentiated neurons in the Neurobasal media containing 2% FCS. 4 µM cytarabine in NBM was added on day-11 of the differentiation to enrich postmitotic neurons. 2/3 of the media were changed every 3-4 days. BrainPhys media (Stem Cell technologies) containing the above mentioned supplements, 3%FCS and 1 µg/ml laminin (Sigma Aldrich) ^85 86^. B27 was depleted for 24h before GR stimulation in BrainPhys based media. Each sample consisted of one well of a 6 well plate.

## Data Availability

All data produced in the present work are contained in the manuscript and PheWAS results via a ShinyApp

https://nprct.shinyapps.io/GRE_PheWAS/

## Acknowledgements

This work was supported by a Joachim-Herz Stiftung Add-on Fellowship for Interdisciplinary Life Science and Canadian Institutes of Health Research Foreign Doctoral Fellowship for Signe Penner-Goeke and the Deutsche Forschungsgemeinschaft, ME4154/1-1, to Laura V. Glaser.

Figures were created with Biorender.com. Figure 4H was adapted from “CRISPR/Cas9 Gene Editing”, by BioRender.com (2022). Retrieved from https://app.biorender.com/biorender-templates Authors from the BeCOME working group are listed in **Table S11**:

## Author Contributions

Conceived and designed project: SPG, EBB

Experimental design: SPG, EBB, ML, SR, SHM, MB, LVG

Collected the data: SPG, EK, DP, AK, MK, JMG, LDH, BW, SS, ML, BeCOME working group

Contributed data or analysis tools: MB, CR, JAK, MJZ, SHM

Performed the analyses: SPG, NK, PK, DP, AK, DC, ML

Critical revision of manuscript: MJZ, JAK

Wrote the paper: SPG, SHM, EBB

## Competing Interests statement

No authors declare a competing interest.

## Notes

### Competing Interest Statement

The authors have declared no competing interest.

### Author Declarations

The Ethics committee of the Ludwigs Maximilians University gave ethical approval for this work and all participants gave an informed consent.

