## Supplemental Figure and Tables Legends and Supplemental Figures for "Assessment of glucocorticoid-induced enhancer activity of eSNP regions using STARR-seq reveals novel molecular mechanisms in psychiatric disorders"

### Supplementary Figures

**Figure S1. Transcriptional response to dex of GR target genes.** Fold changes of expression for three canonical GR-target genes (*FKBP5, SGK1, TSC22D3)* after dex treatment in cells lines as indicated: **A,** U2OS-GR, **B,** U138MG, **C,** induced pluripotent stem cells (IPSCs), **D,** neural precursor cells (NPCs) and E) iNeurons derived from IPSCs from two individuals. Mean expression relative to *GAPDH* ± S.D. is shown, dots depict values for individual data points. One-sided unpaired T-test, * p-value <0.05, ** p-value < 0.01 as compared to expression changes in the control transcript YWHAZ.

**Figure S2. Quality controls of STARR-seq experiments. A,** Principal component analysis of the biological replicates and **B,** Pearson's $R^2$ between biological replicates in the veh and **C,** dex condition for U2OS-GR cells. **D,** Principal component analysis of the biological replicates and **E,** Pearson's $R^2$ between biological replicates in the veh and **F,** dex condition for U138MG cells.

**Figure S3. Enrichment in enhancer sites across tissues.** ChromHMM enrichment of DREs predicted to be located within enhancer regions across different tissues compared to randomly selected matched SNPs. * FDR < 0.05; ** FDR< 0.01; permutation FDR.

**Figure S4. Validation of STARR-seq results using qPCR. A,** Individual fragments of three SNP-DREs were tested for activity in dex and veh conditions to determine the fold activation upon dex treatment. Two of the three variant-DREs displayed significant allele dependent differences in the dex condition (paired, one-sided t-test). **B,** The dex-induced activity for two DRE controls and two STARR-seq identified DREs was tested. Each region showed significant dex-dependent increases in activity when compared to negative control region in the direction observed in the STARR-seq. * p-value < 0.05; ** p-value < 0.01; *** p-value < 0.001. Multiple comparisons test on dex vs. veh normalized Ct values.

**Figure S5. Transcriptional response in response to dex in KO cell lines.** Fold changes of expression for three canonical GR-target genes (*FKBP5, SGK1, TSC22D3)* after dex treatment in WT and **A**, rs12206258 DRE KO **B**, rs35288741 DRE KO cell lines. Note that *TSC22D3* contains a FOXC1 binding site in its promoter, possibly related to the reduced induced of that gene in the KO cells. **C**, Fold changes of transcripts located near rs35288741 in WT and KO cells. NE refers to transcripts not expressed. **D,** Schematic depicting the genomic locus containing an eSNP bin (highlighted orange) in relation to its predicted target gene, *NUAK2* (red box) and surrounding genes. Black box provides a zoomed-in view of the region containing the repressive SNP-DRE rs35288741 with the CRISPR/Cas9-deleted region highlighted in blue. Black dot represents a flanking eSNP not targeted. *p-value < 0.05; ** p-value < 0.01; *** p-value < 0.001, **** p-value < 0.0001, one-sided paired t-test on dex vs. veh normalized Ct values.

**Fig. S6:** Number of PheWAS associations after multiple comparison correction for the top 10 exposures with most associations. Results are categorised by outcome trait category. The y-axis describes the proxy SNP for MR analysis (left side) and associated gene (right side). The x-axis shows the number of PheWAS associations. Points are coloured by functional group.

### Tables

**Table S1. Genomic coordinates (hg19) and allele information for the 3662 eSNPs included in the STARR-seq.**

**Table S2. Dex regulatory elements (DREs) identified in STARR-seq.** Effect sizes for the 508 and 66 DREs identified in the U2OS-GR and U138MG cells for the oligonucleotide harboring the reference and alternative alleles. NS refers to DREs only displaying significant dex-responsive activity in either the reference or alterative allele.

**Table S3.** Motifs significantly enriched within inductive and repressive dex regulatory elements using the Jaspar vertebrate motif database and random human promoters as a background model.

**Table S4. Allele dependent dex-responsive elements (SNP-DREs) identified in STARR-seq.** Effect sizes for the allele-dependent DREs identified in the U2OS-GR and U138MG cells in the dex and veh conditions. NS refers to SNP-DREs only displaying significant allele-dependent differential activity either the dex or veh condition.

**Table S5. Transcripts regulated by DREs and associations with psychiatric disorders.** Number of DREs and SNP-DREs identified as functional using STARR-seq. Associated disorder refers to disorder in which the transcript is differentially expressed in post-mortem tissues of subjects with specific psychiatric disorders.[30]

**Table S6. Mendelian Randomization using SNP-DREs and meta-GWAS on psychopathology.** Cross disorder meta-analysis beta and p-values were derived from the meta-GWAS summary statistics[31] and Mendelian Randomization results for those SNP-DREs showing a significant putative causal effect on psychopathology.

**Table S7.** Unique phenotype with corresponding trait category with significant associations in PheWAS MR analyses.

**Table S8.** Number of PheWAS MR associations across outcome trait categories.

**Table S9.** SNPs used to generate the functional gene score and their respective weightings from the STARR-Seq experiment.

**Table S10.** Primer sequences used for dex-induced gene expression analyses, STARR-seq, and the endogenous and exogenous validation experiments.

**Table S11.** List of authors within the BeCOME working group.

Figure S1

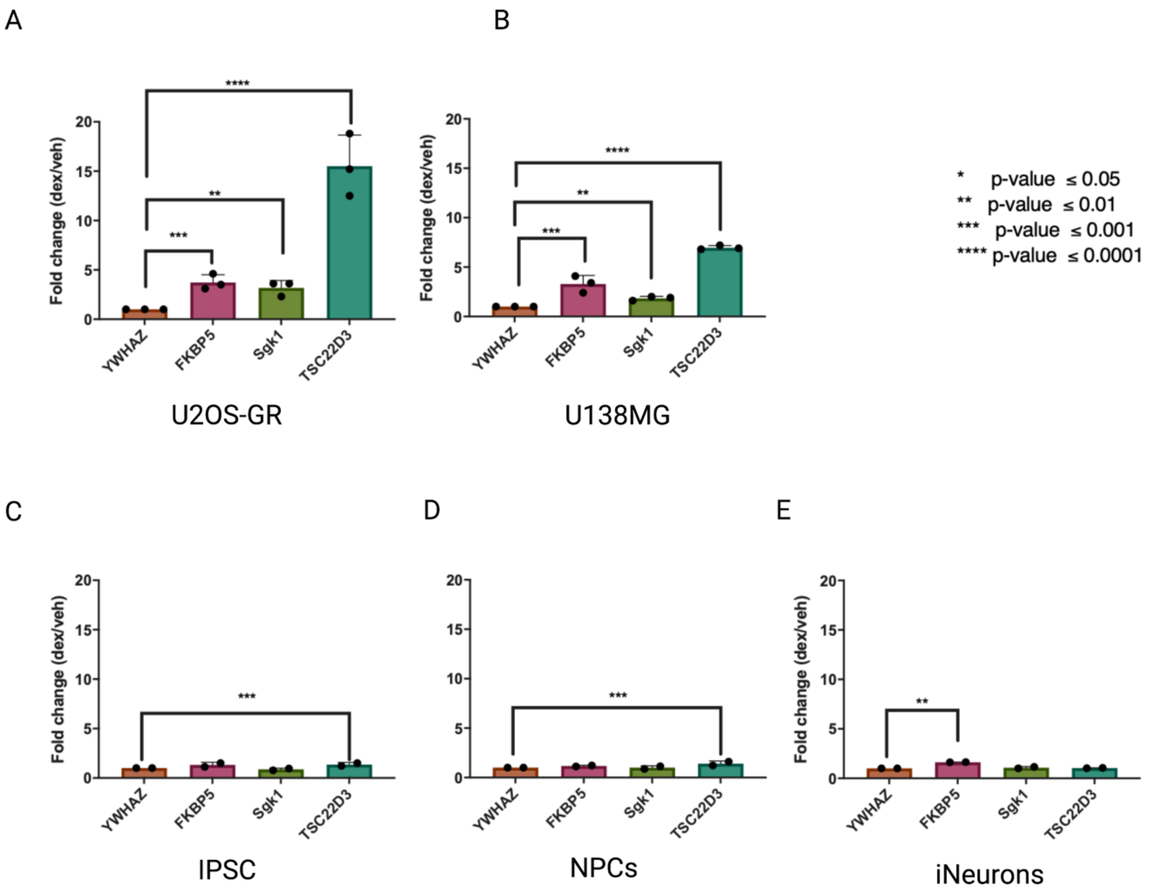
A
B
C
D
E
Fold change (dex/veh)
YWHAZ
FKBP5
Sgk1
TSC22D3
U2OS-GR
U138MG
IPSC
NPCs
iNeurons
* p-value ≤ 0.05
** p-value ≤ 0.01
*** p-value ≤ 0.001
**** p-value ≤ 0.0001

.

Figure S2

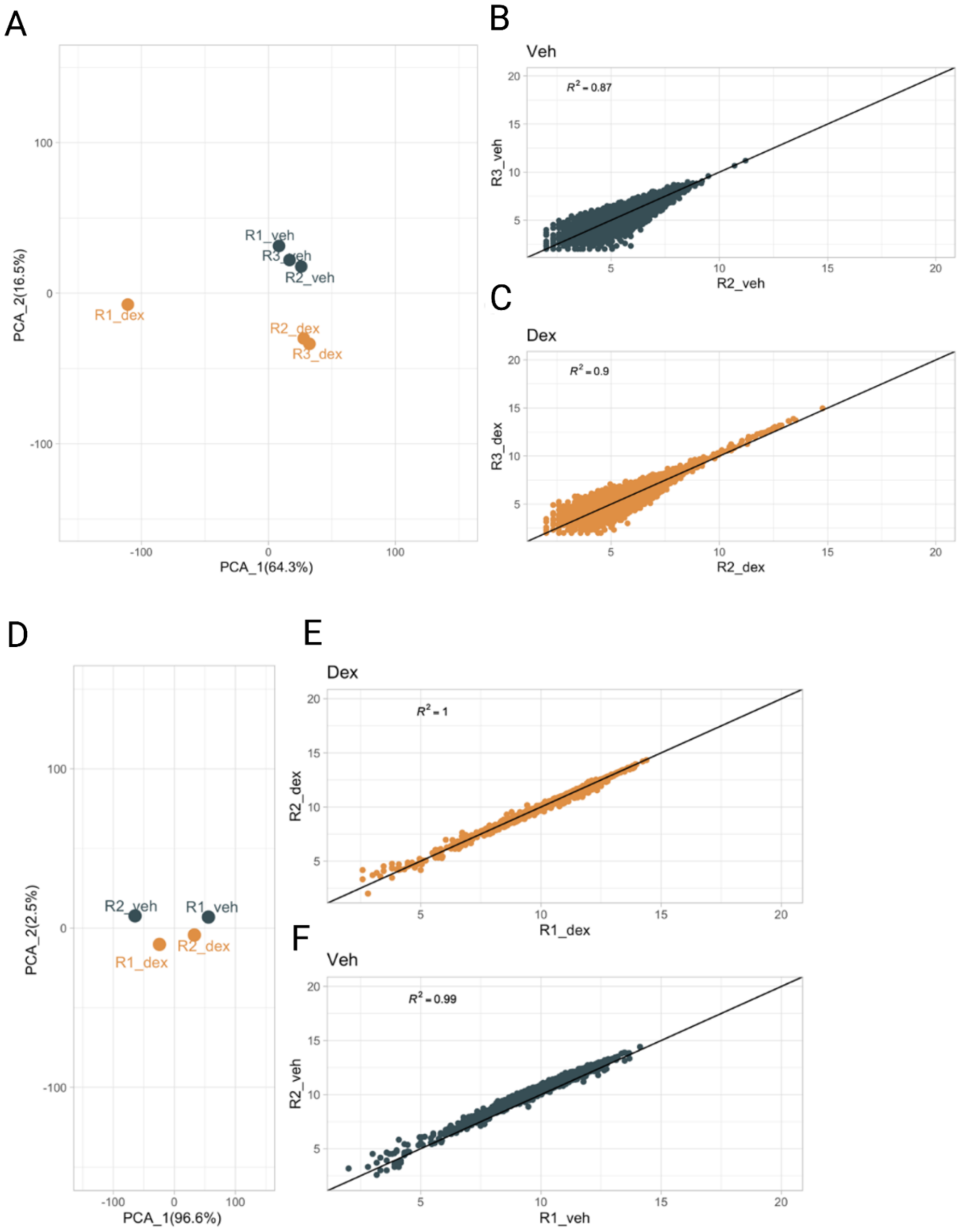
A
B
Veh
$R^2 = 0.87$
R3_veh
R2_veh
C
Dex
$R^2 = 0.9$
R3_dex
R2_dex
PCA_2(16.5%)
PCA_1(64.3%)
R1_veh
R3_veh
R2_veh
R1_dex
R2_dex
R3_dex
D
E
Dex
$R^2 = 1$
R2_dex
R1_dex
F
Veh
$R^2 = 0.99$
R2_veh
R1_veh
PCA_2(2.5%)
PCA_1(96.6%)
R2_veh
R1_veh
R1_dex
R2_dex

Figure S3

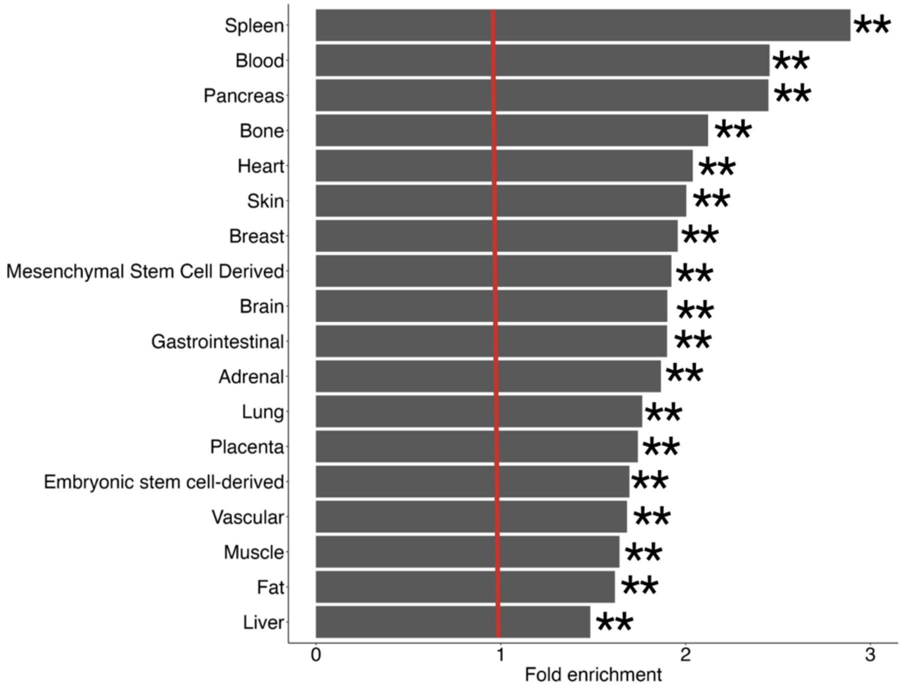
Spleen
Blood
Pancreas
Bone
Heart
Skin
Breast
Mesenchymal Stem Cell Derived
Brain
Gastrointestinal
Adrenal
Lung
Placenta
Embryonic stem cell-derived
Vascular
Muscle
Fat
Liver
0
1
2
3
Fold enrichment

Figure S4

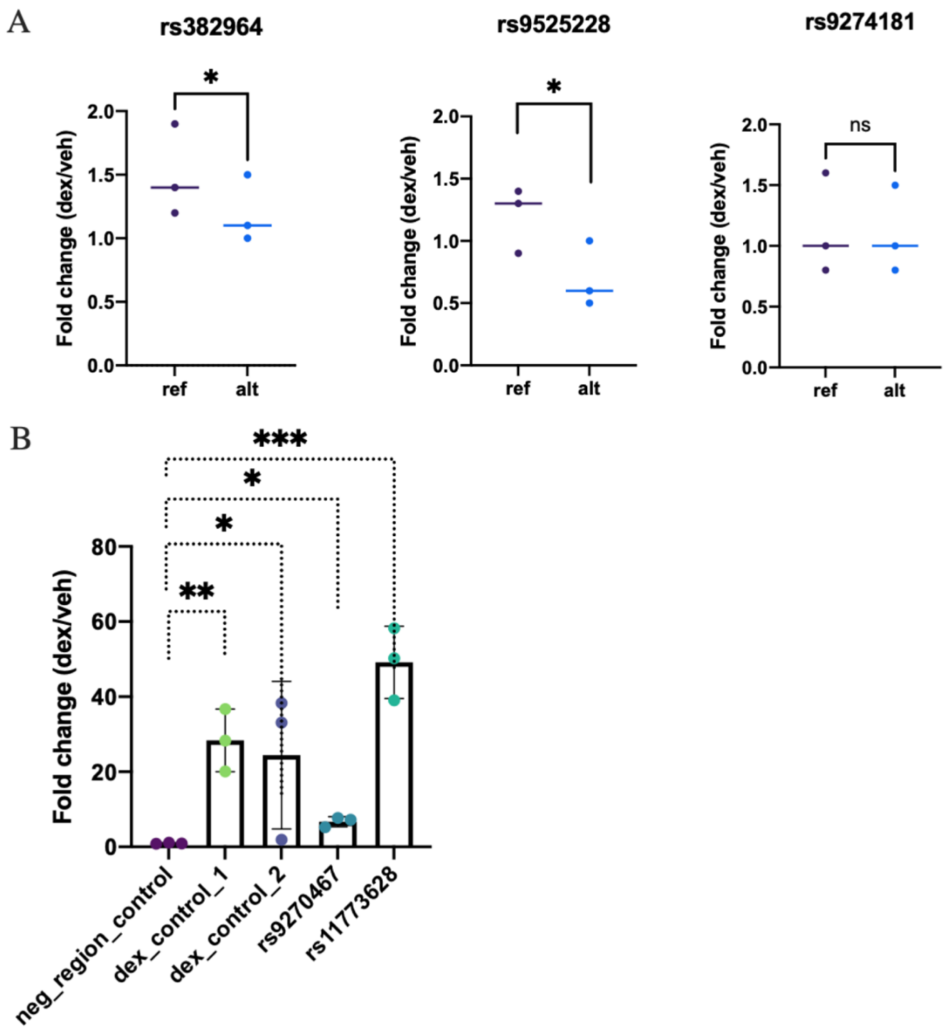
A
rs382964
rs9525228
rs9274181
*
*
ns
Fold change (dex/veh)
2.0
1.5
1.0
0.5
0.0
ref
alt
B
***
*
*
**
80
60
40
20
0
Fold change (dex/veh)
neg_region_control
dex_control_1
dex_control_2
rs9270467
rs11773628

Figure S5

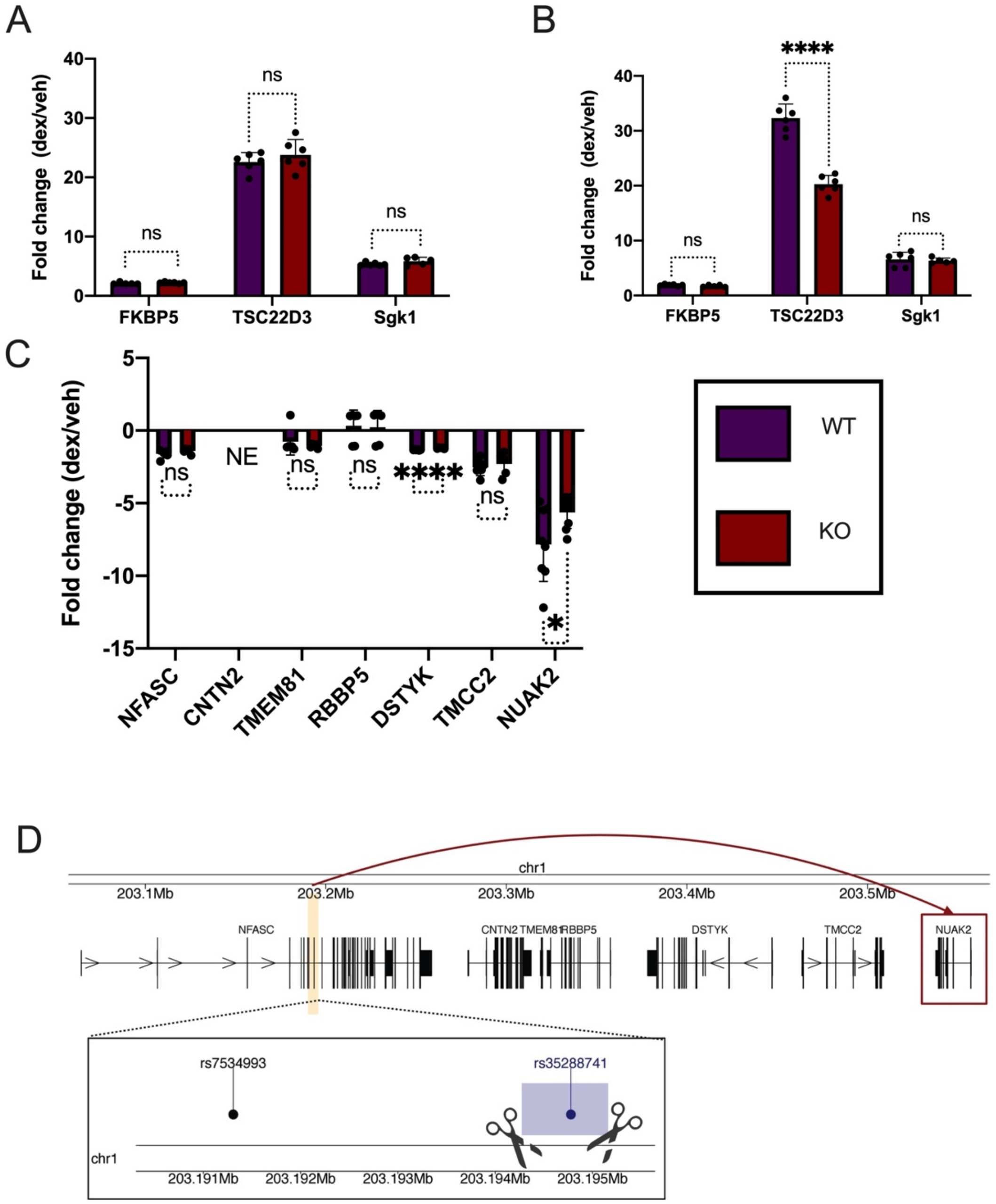

A
Fold change (dex/veh)
40
30
20
10
0
ns
ns
ns
FKBP5
TSC22D3
Sgk1
B
Fold change (dex/veh)
****
ns
ns
C
Fold change (dex/veh)
5
0
-5
-10
-15
NE
ns
ns
ns
****
ns
*
NFASC
CNTN2
TMEM81
RBBP5
DSTYK
TMCC2
NUAK2
WT
KO
D
chr1
203.1Mb
203.2Mb
203.3Mb
203.4Mb
203.5Mb
NFASC
CNTN2 TMEM81 RBBP5
DSTYK
TMCC2
NUAK2
rs7534993
rs35288741
chr1
203.191Mb
203.192Mb
203.193Mb
203.194Mb
203.195Mb

Figure S6

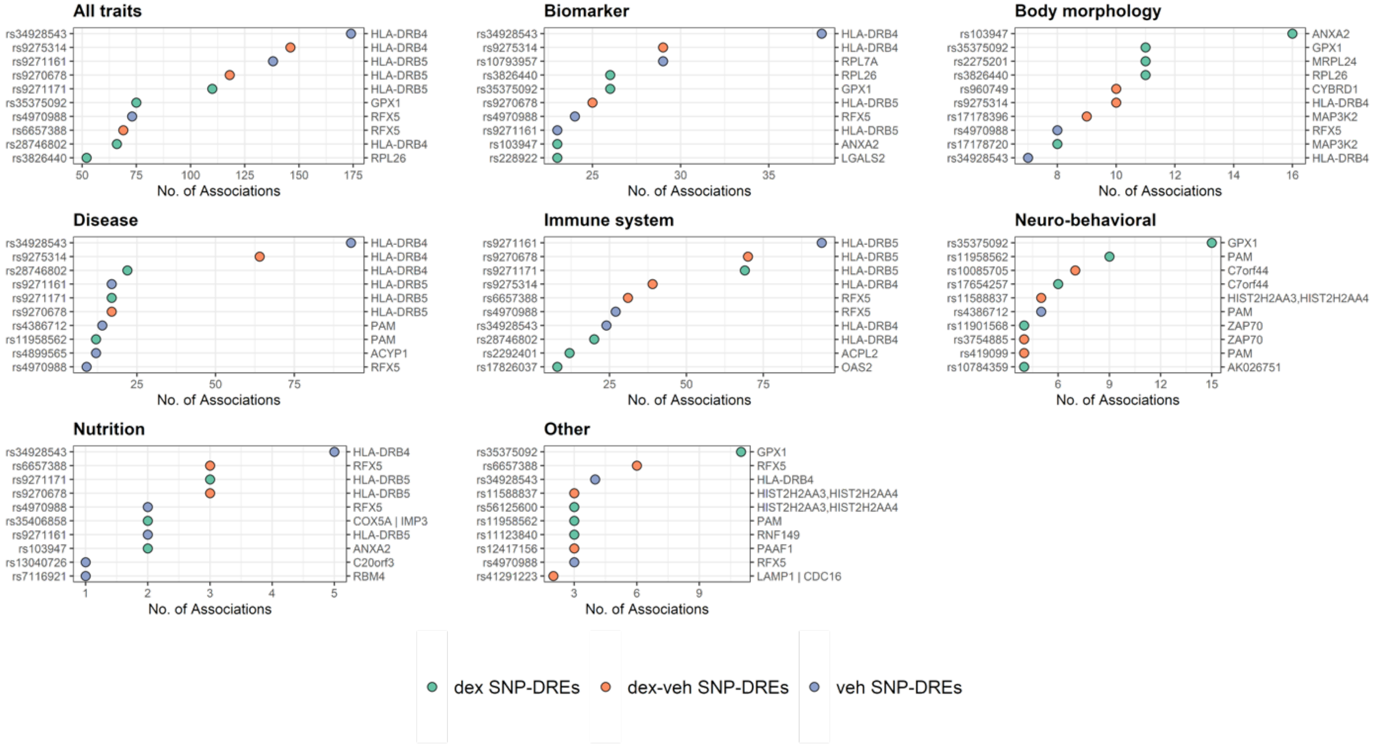
All traits
rs34928543 HLA-DRB4
rs9275314 HLA-DRB4
rs9271161 HLA-DRB5
rs9270678 HLA-DRB5
rs9271171 HLA-DRB5
rs35375092 GPX1
rs4970988 RFX5
rs6657388 RFX5
rs28746802 HLA-DRB4
rs3826440 RPL26
50 75 100 125 150 175
No. of Associations
Biomarker
rs34928543 HLA-DRB4
rs9275314 HLA-DRB4
rs10793957 RPL7A
rs3826440 RPL26
rs35375092 GPX1
rs9270678 HLA-DRB5
rs4970988 RFX5
rs9271161 HLA-DRB5
rs103947 ANXA2
rs228922 LGALS2
25 30 35
No. of Associations
Body morphology
rs103947 ANXA2
rs35375092 GPX1
rs2275201 MRPL24
rs3826440 RPL26
rs960749 CYBRD1
rs9275314 HLA-DRB4
rs17178396 MAP3K2
rs4970988 RFX5
rs17178720 MAP3K2
rs34928543 HLA-DRB4
8 10 12 14 16
No. of Associations
Disease
rs34928543 HLA-DRB4
rs9275314 HLA-DRB4
rs28746802 HLA-DRB4
rs9271161 HLA-DRB5
rs9271171 HLA-DRB5
rs9270678 HLA-DRB5
rs4386712 PAM
rs11958562 PAM
rs4899565 ACYP1
rs4970988 RFX5
25 50 75
No. of Associations
Immune system
rs9271161 HLA-DRB5
rs9270678 HLA-DRB5
rs9271171 HLA-DRB5
rs9275314 HLA-DRB4
rs6657388 RFX5
rs4970988 RFX5
rs34928543 HLA-DRB4
rs28746802 HLA-DRB4
rs2292401 ACPL2
rs17826037 OAS2
25 50 75
No. of Associations
Neuro-behavioral
rs35375092 GPX1
rs11958562 PAM
rs10085705 C7orf44
rs17654257 C7orf44
rs11588837 HIST2H2AA3,HIST2H2AA4
rs4386712 PAM
rs11901568 ZAP70
rs3754885 ZAP70
rs419099 PAM
rs10784359 AK026751
6 9 12 15
No. of Associations
Nutrition
rs34928543 HLA-DRB4
rs6657388 RFX5
rs9271171 HLA-DRB5
rs9270678 HLA-DRB5
rs4970988 RFX5
rs35406858 COX5A | IMP3
rs9271161 HLA-DRB5
rs103947 ANXA2
rs13040726 C20orf3
rs7116921 RBM4
1 2 3 4 5
No. of Associations
Other
rs35375092 GPX1
rs6657388 RFX5
rs34928543 HLA-DRB4
rs11588837 HIST2H2AA3,HIST2H2AA4
rs56125600 HIST2H2AA3,HIST2H2AA4
rs11958562 PAM
rs11123840 RNF149
rs12417156 PAAF1
rs4970988 RFX5
rs41291223 LAMP1 | CDC16
3 6 9
No. of Associations
dex SNP-DREs
dex-veh SNP-DREs
veh SNP-DREs
